# Differential T cell reactivity to seasonal coronaviruses and SARS-CoV-2 in community and health care workers

**DOI:** 10.1101/2021.01.12.21249683

**Authors:** Ricardo da Silva Antunes, Suresh Pallikkuth, Erin Williams, Esther Dawen Yu, Jose Mateus, Lorenzo Quiambao, Eric Wang, Stephen A. Rawlings, Daniel Stadlbauer, Kaijun Jiang, Fatima Amanat, David Arnold, David Andrews, Irma Fuego, Jennifer M. Dan, Alba Grifoni, Daniela Weiskopf, Florian Krammer, Shane Crotty, Michael E. Hoffer, Savita G. Pahwa, Alessandro Sette

## Abstract

Herein we measured CD4^+^ T cell responses against common cold corona (CCC) viruses and SARS-CoV-2 in high-risk health care workers (HCW) and community controls. We observed higher levels of CCC reactive T cells in SARS-CoV-2 seronegative HCW compared to community donors, consistent with potential higher occupational exposure of HCW to CCC. We further show that SARS-CoV-2 reactivity of seronegative HCW was higher than community controls and correlation between CCC and SARS-CoV-2 responses is consistent with cross-reactivity and not associated with recent in vivo activation. Surprisingly, CCC reactivity was decreased in SARS-CoV-2 infected HCW, suggesting that exposure to SARS-CoV-2 might interfere with CCC responses, either directly or indirectly. This result was unexpected, but consistently detected in independent cohorts derived from Miami and San Diego.

## Introduction

Coronavirus Disease (COVID-19) caused by the novel zoonotic pathogen severe acute respiratory syndrome coronavirus 2 (SARS-CoV-2) is responsible for a global pandemic and accounting for 1.8 million deaths and over 85 million cases worldwide and over 21 million cases and 360 thousand deaths in United States alone as of January 2021 (Covid.cdc.gov; https://coronavirus.jhu.edu/map.html), as well as dramatic economic and social impact.

Healthcare workers (HCW) that provide frontline care are at increased risk of infection due to frequent close and prolonged exposure to patients with SARS-CoV-2 (1). SARS-CoV-2 infection rates among HCW are still largely undetermined and highly variable depending on the geographical and temporal distribution among other factors (2-5) but higher prevalence has been documented during periods of upsurge (6, 7). Still, only a minority have developed mild to severe disease manifestations and the majority have remained seronegative for SARS-CoV-2 antibodies despite having close contact with SARS-CoV-2 infected patients (2-4, 8, 9). Interestingly, a relatively low prevalence of COVID-19 in HCW from diverse specialties at the University of Miami in South Florida has been reported (https://coronavirus.miami.edu/dashboard/) in an area with a very high community prevalence of COVID-19 cases which could suggest less susceptibility to infection in this particular cohort.

Robust T cell immunity has been consistently reported in multiple studies in asymptomatic, acute, and convalescent COVID-19 individuals (10-12). Furthermore, we and others have previously reported significant pre-existing immune memory responses to SARS-CoV-2 sequences in unexposed subjects (10, 12-15). Here, we aimed to characterize preexisting SARS-CoV-2 T cell responses in this HCW cohort.

Due to close contact with patients, HCW are particularly prone to exposure to respiratory pathogens such as human coronaviruses (HCoVs) and particularly to endemic “common cold” corona virus (CCC) (16-18) (http://www.cdc.gov/niosh/topics/healthcare/infectious.html). <u>Human</u> CCC (comprising either the alphacoronaviruses 223E and NL63, or the betacoronaviruses OC43 and HKU1) are seasonal endemic circulating viruses that cause only mild upper and lower respiratory infections. They are globally distributed with higher incidences in winter months. Little is known about their pattern of infection, transmission rates, or duration of immunity (19-21). As expected, on the basis of their common phylogeny, CCC share varying degrees of sequence homology with SARS-CoV-2 and we and others have shown that cross-reactive CD4^+^ T cell memory responses against SARS-CoV-2 can be detected in unexposed donors (13, 22, 23). While detection of pre-existing immunity to CCC has mainly been described in studies focusing on T cell responses, potential antibody-based cross-reactivity or neutralizing activity has also been suggested (24-27).

However, it is still unclear how pre-existing immunity impacts disease severity or clinical outcome after SARS-CoV-2 exposure (28, 29) and if this could translate into a protective effect. While some studies suggest this could be the case (25, 30, 31), and exposure to CCC concomitantly results in a faster response of pre-existing memory cells to control SARS-CoV-2 infection, it cannot be excluded that CCC cross-reactivity could contribute to drive COVID-19 immunopathogenesis (32). Thus, it is important to study differences in CCC reactivity and pre-existing immunity in different cohorts, particularly HCW.

## Results

### Characteristics of the donor cohorts investigated

Five different cohorts of subjects were enrolled in the study (**Table 1**). Three cohorts were recruited in the Miami metropolitan area and two cohorts were recruited in the San Diego metropolitan area. Two cohorts from Miami encompassed high-risk HCW (composed of individuals from the fields of Otolaryngology, Anesthesiology, Emergency Medicine and Ophthalmology), further classified as sero<u>N</u>egative Healthcare Workers (NHCW) or Antibody or PCR <u>P</u>ositive Healthcare Workers (PHCW); the third Miami cohort designated Shelter In Place (SIP) community volunteers who were all seronegative and with no exposure to known infected persons. The two additional cohorts were asymptomatic unexposed and sero<u>N</u>egative donors from San Diego (NSD), and COVID-19 seropositive subjects also from the San Diego region (COVID-19SD).

**Table 1.** Description of donor cohort characteristics and demographics

| Cohort Name | NHCW<sup>+</sup> | PHCW<sup>++</sup> | SIP<sup>+++</sup> | NSD<sup>++++</sup> | COVID-19 SD<sup>++++</sup> |
| --- | --- | --- | --- | --- | --- |
| **Geographical Location** | Miami | Miami | Miami | San Diego | San Diego |
| **Number of donors** | 32 | 26 | 33 | 15 | 10 |
| Gender (M/F) | (17, 15) | (13, 13) | (16, 17) | (7, 8) | (3, 7) |
| Mean age (years) | 41 | 38 | 41 | 41 | 32 |
| **Sample collection date** | Apr-Jun 2020 | Aug 2020 | Jun 2020 | Mar-Jun 2020 | Apr-Jul 2020 |
| **SARS-CoV-2 status** | Ab(-) | Ab(+) or PCR(+) | Ab(-) | Ab(-) | Ab(+) |
| **SARS-CoV-2 PCR (n (%))** |  |  |  |  |  |
| Positive | 0 (0) | 22 (84.6) | 0 (0) | 0 (0) | 7 (70) |
| Unknown | - | 4 (15.4) | - | - | 3 (30) |
| **Antibody response (n (%))** |  |  |  |  |  |
| Positive | 0 (0) | 23 (88.5) | 0 (0) | 0 (0) | 10 (100) |
| Negative | 32 (100) | 3 (11.5) | 33 (100) | 15 (100) | 0 (0) |
| **Interval exposure to sample collection (days)** |  |  |  |  |  |
| Range | - | 20-145 | - | - | 43-140 |
| Median | - | 44 | - | - | 92 |
| **Symptoms (n (%))** |  |  |  |  |  |
| Asymptomatic | 32 (100) | 7 (26.9) | 33 (100) | 15 (100) | 0 (0) |
| Mild | - | 19 (73) | - | - | 8 (80) |
| Moderate | - | 0 (0) | - | - | 1 (10) |
| Severe | - | 0 (0) | - | - | 1 (10) |
| **Medical speciality (n (%))** |  |  |  |  |  |
| Otolaryngology | 17 (53.1) | 6 (23.1) | - | - | - |
| Anesthesiology | 5 (15.6) | 3 (11.5) | - | - | - |
| Emergency Medicine | 5 (15.6) | 5 (19.2) | - | - | - |
| Ophthalmology | 2 (6.3) | 2 (7.7) | - | - | - |
| Other (internal medicine, surgery, etc.) | 3 (9.4) | 10 (38.5) | - | - | - |

Summary of donor characteristics of the Miami and San Diego cohorts.
+ indicates seronegative high-risk health care workers from Miami; ++ indicates antibody (Ab) /PCR positive high-risk health care workers from Miami; +++ indicates seronegative community donors with no patient exposure from Miami; ++++ indicates seronegative unexposed donors from San Diego; +++++ indicates seropositive donors from San Diego.

The number of donors in each cohort ranged from 10 to 32 subjects (median 26), and mean age ranged from 32 to 41 (median 41) years. All samples were collected in the March to August 2020 period. All subjects were assigned to positive or negative SARS-CoV-2 categories on the basis of PCR and/or serological tests. For the two seropositive cohorts, the intervals from exposure to sample collection were 20-145 days (median 44) for Miami, and 43-140 (median 92) for San Diego. Most donors were associated with mild cases. For HCW cohorts, the medical specialty of the different subjects is detailed in **Table 1**.

### Serological analysis of the different donor cohorts

Serum samples for all five donor cohorts were tested for SARS-CoV-2 using the enzyme linked immunosorbent assay (ELISA) (see methods for detail). For the SIP cohort, a N-antigen ELISA assay for IgG and IgM that was purely qualitative was performed. All donors had undetectable levels of antibodies.

For the NHCW, PHCW, NSD and COVID-19SD the serologic test used was based on a two-step ELISA with an initial screening for antibodies to the receptor binding domain of the SARS-CoV-2 spike protein followed by titration using full length trimeric stabilized spike as substrate. Only samples that produced a positive signal in both assays were counted as true positives. The results are shown in **Figure 1A**. Significant SARS-CoV-2 titers were detected in almost all cases of individuals in the HCW cohort with COVID-19 disease from Miami (23/26). Interestingly the 4 donors with the lowest titers and 2 out of 3 donors without detectable titers, despite being PCR positive for SARS-CoV-2, were asymptomatic or did not report experiencing any COVID-19 disease symptoms. Conversely, the seronegative cohorts from Miami (NHCW) had undetectable titers or below the limit of detection. Likewise, all COVID-19SD had significant SARS-CoV-2 titers, while none of the NSD donors was seropositive for SARS-CoV-2 spike antibodies.

**Figure 1.**
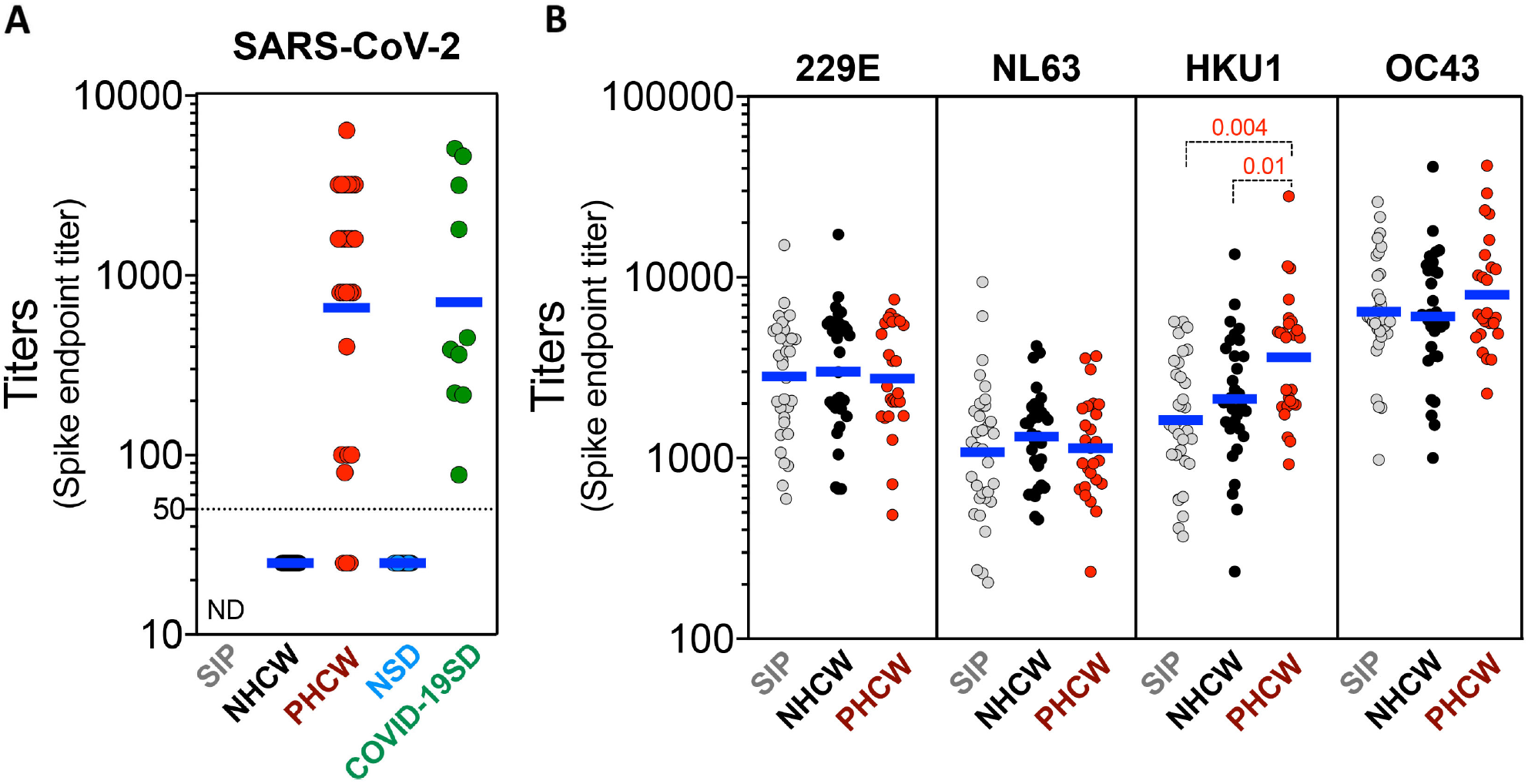
SARS-CoV-2 and CCC viruses serological reactivity for the donor cohort. (A) Serum ELISA titers to SARS-CoV-2 spike RBD protein. “ND” = Not Determined. (B) Serum ELISA titers to CCC viruses (HCoV-229E, HCoV-NL63, HCoV-HKU1 and HCoV-OC43) spike protein. Non-parametric Kruskal-Wallis multiple comparison test was applied for each individual CCC strain. Geometric mean titers are indicated and p values are shown for the statistical significant comparisons. “SIP” = Shelter In Place community volunteers (n=33). “NHCW” = SeroNegative Health Care Workers (n=31). “PHCW” = Antibody or PCR Positive Health Care Workers (n=26). “NSD” = SeroNegative San Diego (n=15). “COVID-19” = Seropositive San Diego (n=10). Dotted line indicates limit of detection (1:50).

In parallel, seropositivity for the spike proteins of the four seasonal common cold coronaviruses (CCC; 229E, NL63, HKU1 and OC43), was also determined in the three donor cohorts from Miami (**Figure 1B)**. All donors had detectable titers and variable reactivity for each of the CCC strains. Antibody reactivity against NL63 was the lowest whilst OC43 was the highest across all the cohorts. Titers for the HKU1 betacoronavirus, were significantly higher for the PHCW cohort, consistent with the reported back-boosting effect to HKU1 and OC43 spike in COVID-19 cases (26). Taken together, these results define the three cohorts investigated in terms of serological reactivity to SARS-CoV-2 and CCC. The results are further consistent with the majority of the general population having detectable responses for the CCC viruses (19, 20, 33).

In conclusion, these data define the serological status of the donor cohorts for which the T cell reactivity was investigated.

### CD4^+^ T cell reactivity against CCC is higher in NHCW compared to SIP and PHCW

To test the various Miami cohorts for CD4^+^ T cell reactivity, we performed Activation Induced Marker (AIM) assays (34, 35). AIM is a commonly used methodology to detect antigen-specific cells with high specificity and sensitivity (34-36) previously utilized to characterize viral responses (37, 38) and particularly SARS-CoV-2 CD4^+^ T cell responses, utilizing the OX40 (CD134) and 4-1BB (CD137) markers (12, 14, 15, 39). To test the hypothesis that NHCW would have higher CD4^+^ T cell reactivity to CCC viruses, we also generated sets of predicted dominant Class II-restricted T cell peptides, for each of the four CCCs. The reference genome of each virus was scanned using a previously described algorithm to predict promiscuous dominant T cell epitopes (40, 41), and a total of 225 to 294 epitopes were predicted for the different viruses (229E, NL63, HKU1 and OC43). This epitope prediction strategy was previously applied in multiple studies (34, 36, 42) and was envisioned to capture the top 50% of the predicted response.

For assessing CCC responses in the different cohorts, the cross-reactivity between CCC and SARS-CoV-2 had to be accounted for to prevent confounding the analysis, especially in PHCW who were exposed to both CCC and SARS-CoV-2 viruses. Our previous study indicated that CCC-SARS-CoV-2 cross-reactive epitopes shared 67% or higher homology (15). Accordingly, to derive CCC epitope pools with reduced SARS-CoV-2 cross-reactivity, any CCC epitope sharing 67% or more homology with SARS-CoV-2 sequences were removed to assess specific and not cross-reactive CD4^+^ T cell responses. Depleting the peptides with high sequence homology resulted in the generation of CCC pools containing 205 to 272 different predicted epitopes (**Supplementary Table 1**).

The CD4^+^ T cell reactivity to the 229E, NL63, HKU1 and OC43 viruses was higher in the NHCW cohort, as compared to the SIP cohort (**Figures 2a-b** show absolute magnitude and stimulation index (SI) plots). This difference was most pronounced for NL63 and least pronounced for HKU1 (p values ranged from 0.02 to 0.0004 by the Kruskal-Wallis test).

**Figure 2.**
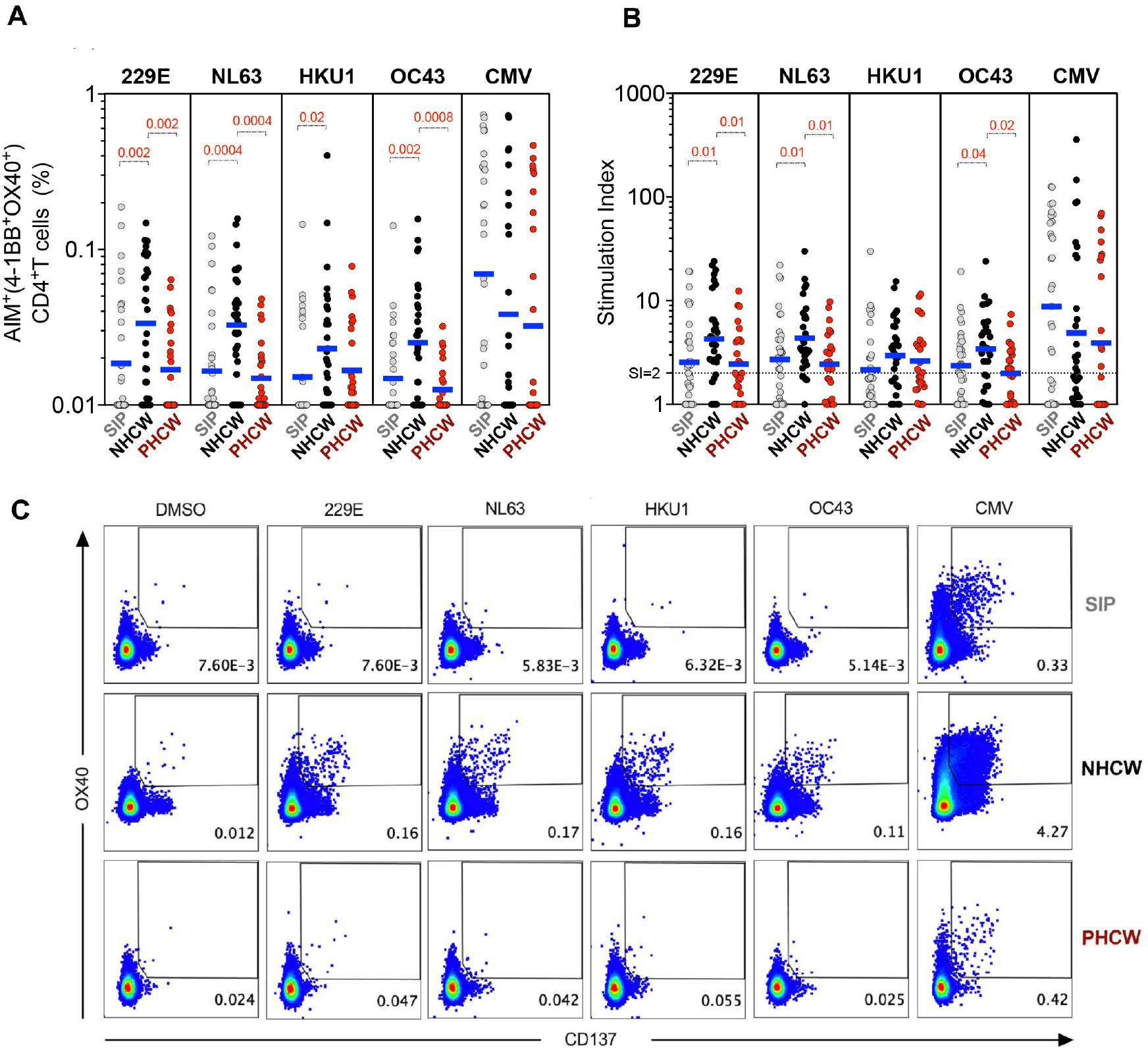
CD4+ T cell immune responses to CCC epitopes from Miami are higher in NHCW. CCC-specific CD4+ T cells (HCoV-229E, HCoV-NL63, HCoV-HKU1 and HCoV-OC43) and ubiquitous control CMV-specific CD4+ T cells were measured as percentage of AIM+ (OX40+CD137+) CD4+ T cells after stimulation of PBMCs with CCC and CMV peptide pools. (A) Data background subtracted or (B) stimulation index (SI) against DMSO negative control are shown with geometric mean for the 3 different groups. Non-parametric Kruskal-Wallis multiple comparison test was applied for each individual CCC strain and CMV. P values are shown for the statistical significant comparisons. “SIP” = Shelter In Place community volunteers (n=33). “NHCW” = SeroNegative Health Care Workers (n=31). “PHCW” = Antibody or PCR Positive Health Care Workers (n=26). (C) Representative FACS plots, gated on total CD4+ T cells for the 4 CCC in addition to the DMSO and CMV across all the cohorts. Cell frequency for AIM+ cells in the several conditions is indicated.

By contrast, NHCW CD4^+^ T cell reactivity was significantly higher compared to PHCW against 229E, NL63 and OC43 (p values ranging from 0.002 to 0.0004). For HKU1 there was a trend toward higher responses (p=0.17). No difference was noted with a control MP composed of epitopes derived from the unrelated ubiquitous cytomegalovirus (CMV) pathogen (43). Representative flow cytometry plots with CCC-specific and CMV CD4^+^ T cell responses are shown in **Figure 2c**.

### CD4^+^ T cell reactivity against CCC is higher in unexposed compared to COVID-19 donors in an independent cohort

To validate these results further, we assessed CCC responses in two additional cohorts recruited in the San Diego region, selected on the basis of being asymptomatic and seronegative (NSD) or symptomatic and seropositive (COVID-19SD) for SARS-CoV-2 infection. (**Table 1**). Both cohorts were recruited between March and July of 2020, similar to the Miami cohort. COVID-19 cases were predominantly mild symptomatic cases, and PBMC samples collection times after exposure ranged from 43 to 140 days (Median= 92 days) PSO (**Table 1**).

The 229E, NL63, HKU1 and OC43 epitope pools displayed higher CD4^+^ T cell reactivity in the unexposed donors, as compared to the COVID-19 diagnosed donors, regardless of whether absolute numbers or SI values were considered (**Figures 3a-b)**. No differences between groups were observed in the responses against the CMV control MP. These results indicate that healthy unexposed donors demonstrate higher CD4^+^ T cell reactivity against CCC than COVID-19 donors.

**Figure 3.**
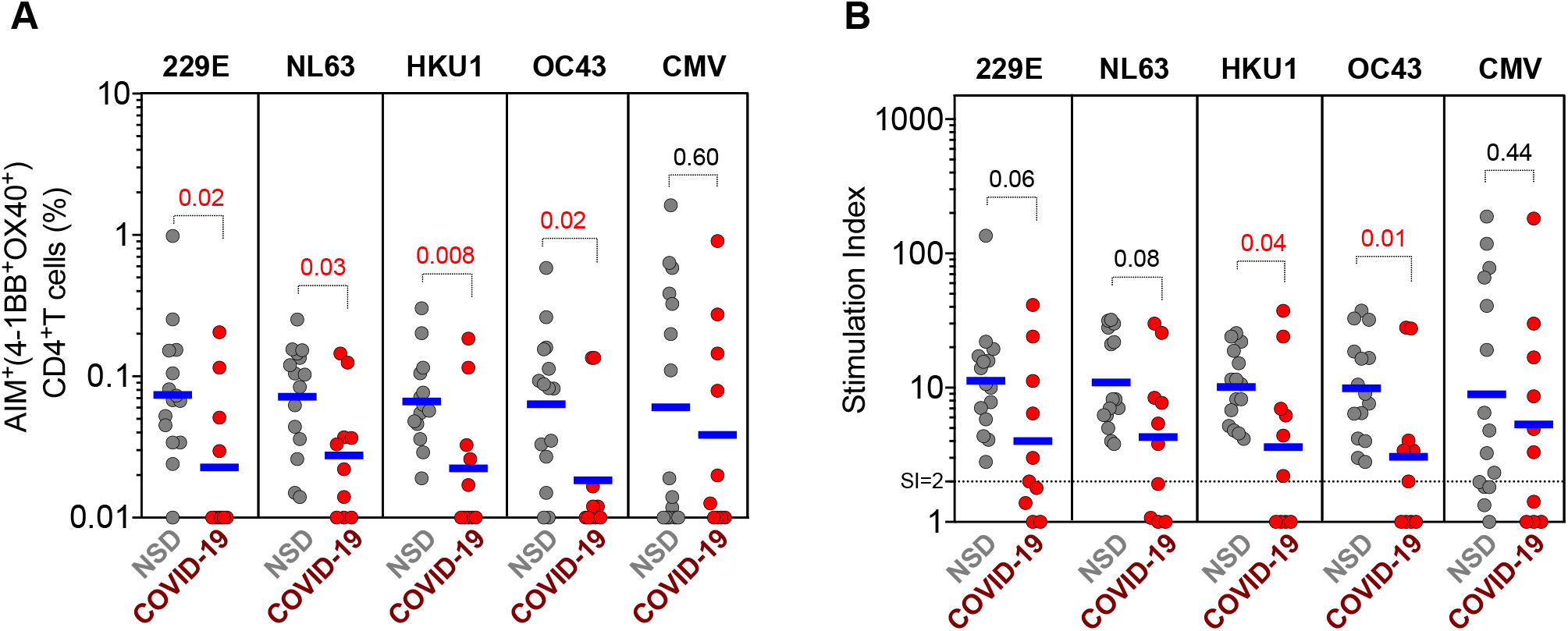
Reactivity of CD4+ T cells against CCC epitopes in an independent cohort from San Diego. CCC-specific CD4+ T cells (HCoV-229E, HCoV-NL63, HCoV-HKU1 and HCoV-OC43) and ubiquitous control CMV-specific CD4+ T cells were measured as percentage of AIM+ (OX40+CD137+) CD4+ T cells after stimulation of PBMCs with CCC and CMV peptide pools. (A) Data background subtracted or (B) stimulation index (SI) against DMSO negative control are shown with geometric mean for the 2 different groups. Samples were from unexposed seronegative donors (“NSD”, n=15) and recovered COVID-19 patients (“COVID-19”, n=10). Statistical comparisons across cohorts were performed with the Mann-Whitney test. P values are shown with p<0.05 defined as statistical significant.

### CD4^+^ T cell reactivity to SARS-CoV-2 S and CD4R MPs

Next, we tested the various cohorts from the Miami area for SARS-CoV-2 CD4^+^ T cell reactivity, using the AIM assay and previously described MPs, one encompassing overlapping peptides spanning the entire sequence of the SARS-CoV-2 spike protein (S), and one encompassing predicted CD4^+^ T cell epitopes from the remainder of the genome (CD4R) (12, 44) (**Supplementary Table 1**). The results are shown in **Figure 4a**, which depict CD4^+^ T cell responses in the various cohorts plotted as background subtracted data. A representative flow cytometry AIM^+^ gating is shown in **Supplementary Figure 1**. CD4^+^ T cell responses plotted as stimulation index are also shown in **Figure 4b**. The graphs display the responses against the two different epitope pools separately, and summed together.

**Figure 4.**
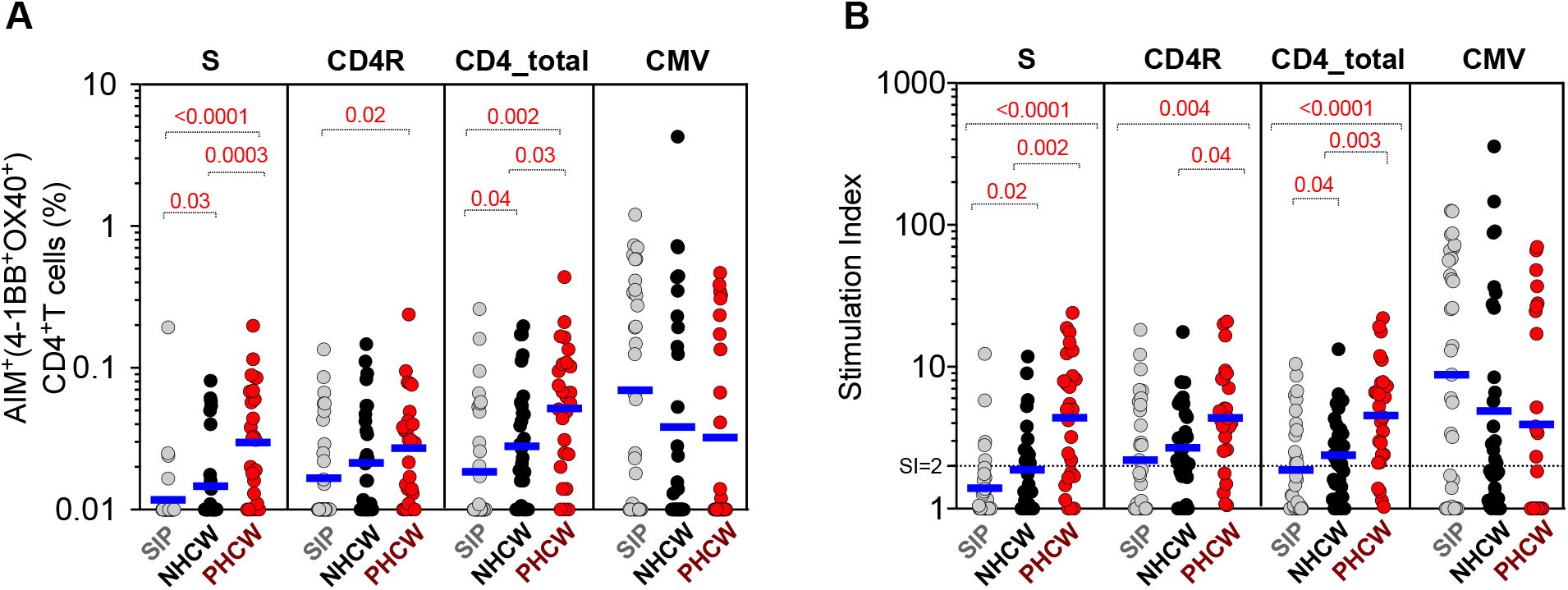
CD4+ T cell response to SARS-CoV-2 epitopes highest in PHCW and lowest in SIP. SARS-CoV-2-specific CD4+ T cells were measured as percentage of AIM+ (OX40+CD137+) CD4+ T cells after stimulation of PBMCs with peptide pools encompassing spike (“S”) or representing all the proteome without spike (“CD4R”). Graphs show data for specific responses against S, CD4R or the combination of both (CD4-total) and against CMV as a control, and plotted as (A) background subtracted or (B) as stimulation index (SI) against DMSO negative control. Geometric mean for the 3 different groups is shown. Non-parametric Kruskal-Wallis multiple comparison test was applied. P values are shown for the statistical significant comparisons. “SIP” = Shelter In Place community volunteers (n=33). “NHCW” = SeroNegative Health Care Workers (n=31). “PHCW” = Antibody or PCR Positive Health Care Workers (n=26).

CD4^+^ T cell responses from PHCW cohort were highest, in accordance with their recent exposure to SARS-CoV-2, followed by responses measured in the NHCW and then the SIP cohort. More specifically, the total CD4^+^ T cell reactivity of the PHCW cohort to the SARS-CoV-2 pools was significantly higher than both NHCW (p =0.03 and p =0.003 by the Kruskal-Wallis test for absolute and SI readouts, respectively) and SIP (p =0.002 and p <0.0001 by the Kruskal-Wallis test for absolute and SI readouts, respectively). Of further interest, the total CD4^+^ T cell reactivity of NHCW was also higher than that observed in the SIP cohort (p=0.04 for both absolute and SI readouts). No difference was noted in the case of the CMV MP. Analysis of the expression of the CCR7 and CD45RA memory markers confirmed that the CD4^+^ T cell reactivity in all three cohorts was mediated by memory T cell subsets (**Supplementary Figure 2)**.

The relatively higher reactivity to SARS-CoV-2 sequences observed in NHCW, may result from higher levels of CCC reactivity, resulting in higher levels of responses cross-reactive with SARS-CoV-2 sequences. Alternatively, it cannot be excluded that some of the NHCW might have been previously infected, despite being seronegative at the time of the blood draw.

### SARS-CoV-2 reactivity in NHCW is not likely due to resolved SARS-CoV-2 infections in absence of seroconversion

As shown in **Figure 4** the PHCW had a higher CD4^+^ T cell reactivity than the SIP and NHCW, in particular against the Spike protein (S). This data is compatible with the notion that the CD4^+^ T cell reactivity to SARS-CoV-2 sequences detected in SIP and NHCW is due to CCC exposure (15, 23, 28), while the reactivity detected in PHCW is due to SARS-CoV-2 exposure.

To address this issue further, we analyzed the AIM^+^ CD4^+^ T cells detected in the three Miami cohorts upon *ex vivo* stimulation with the S and CD4R pools for expression of the HLA-DR/CD38 markers, which have been found increased in donors from mild to acute SARS-CoV-2 infection, and therefore to be associated with recent *in vivo* activation (11, 45, 46). The data shown in **Figure 5**, demonstrates that the CD4^+^ T cell reactivity to SARS-CoV-2 peptides is associated with an increased fraction of recently activated T cells in the case of the PHCW cohort, as compared to the NHCW or SIP cohorts. These results are compatible with recent SARS-CoV-2 infection of the PHCW cohort but not for the NHCW or SIP cohorts. No difference was detected in the case of the epitope pool derived from the control ubiquitous antigen CMV. In conclusion, the analysis of HLA DR/CD38 markers results are most consistent with the higher reactivity in NHCW not being due to SARS-CoV-2 infection.

**Figure 5.**
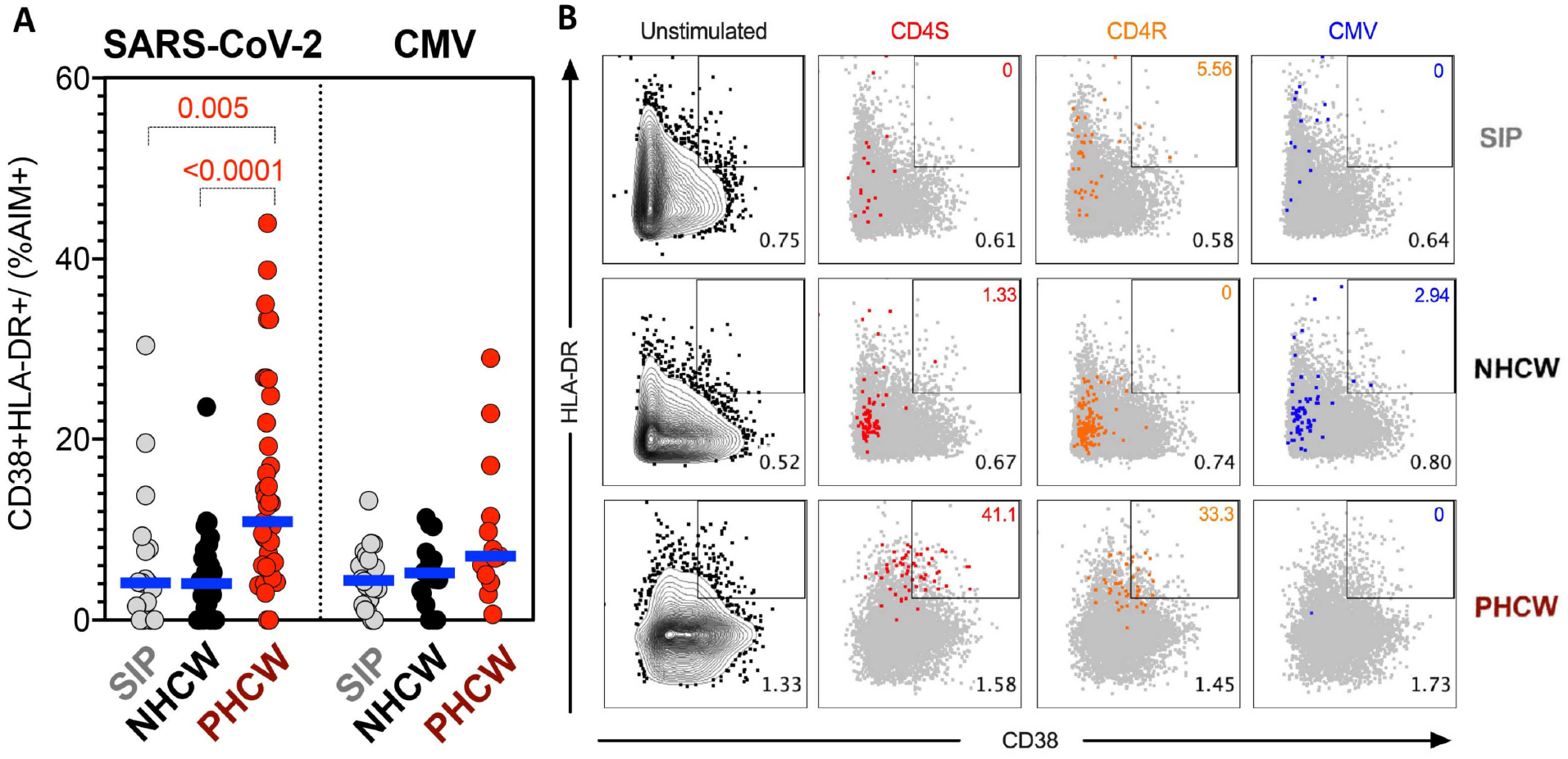
Highest PHCW reactivity in CD4+ T cell responses associated with recent infection. (A) Recently activated SARS-CoV-2-specific CD4+ T cells were measured as percentage of CD38+/HLA-DR+ cells in AIM+ (OX40+CD137+) CD4+ T cells after stimulation of PBMCs with peptide pools encompassing a spike only (“S”) MP and MP representing all the proteome without spike (“CD4R”). Graphs show data for specific responses against SARS-CoV-2 (both “S” and “CD4R”) the ubiquitous pathogen CMV of responses with SI>2. Each dot represents the response of an individual subject to an individual pool. Geometric mean for the 3 different groups is shown. Non-parametric Kruskal-Wallis multiple comparison test was applied. P values are shown for the statistical significant comparisons. “SIP” = Shelter In Place community volunteers (n=20). “NHCW” = SeroNegative Health Care Workers (n=33). “PHCW” = Antibody or PCR Positive Health Care Workers (n=39). (B) Representative FACS plots of HLA-DR/CD38+ cells in AIM+ (OX40+CD137+) CD4+ T cells (colored) overlapped with total HLA-DR/CD38 expression (grey) for all the cohorts in the different unstimulated or stimulated conditions. Cell frequency of HLA-DR/CD38+ in AIM+ cells or total CD4+ T cells is indicated on the top and bottom right corner respectively.

Having measured CCC specific responses we further examined responses on a donor-by-donor basis, and asked whether donors with high CCC CD4^+^ T cell reactivity also have high SARS-CoV-2 CD4^+^ T cell reactivity. A strong correlation was detected between total CD4^+^ T cell responses to CCC and SARS-CoV-2 **(Supplementary Figure 3)** in all the cohorts and for all CCC strains, but less pronounced for OC43 in PHCW (significant p values ranged from 0.0125 to <0.0001). These results suggest that SARS-CoV-2 reactivity is likely due to cross-reactive CCC CD4^+^ T cell responses in both seronegative cohorts. Furthermore, responses to CCC and SARS-CoV-2 were also correlated in the PHCW cohort while no correlation was observed between SARS-CoV-2 and CMV responses **(Supplementary Figure 3)**.

### CD8^+^ T cell reactivity to SARS-CoV-2 epitopes

Finally, we measured CD8^+^ T cell reactivity to SARS-CoV-2 epitopes in the various cohorts. For this purpose, we used the AIM assay we previously utilized to characterize SARS-CoV-2 CD8^+^ T cell responses (12, 14), utilizing the CD69 and 4-1BB markers. For antigen stimulation, we utilized the previously described pool of overlapping peptides spanning the S antigen, and two MPs containing SARS-CoV-2 predicted HLA binders for the 12 most common HLA A and B alleles (CD8A and CD8B MPs) (**Supplementary Table 1)**.(44)

Representative flow cytometry plots are shown in **Supplementary Figure 4. Figure 6A** shows CD8^+^ T cell responses plotted as background subtracted data, or plotted as stimulation index as shown in **Figure 6B**. The graphs show the responses against the S pool, the two different CD8A and CD8B epitope summed together and the control CMV pool.

**Figure 6.**
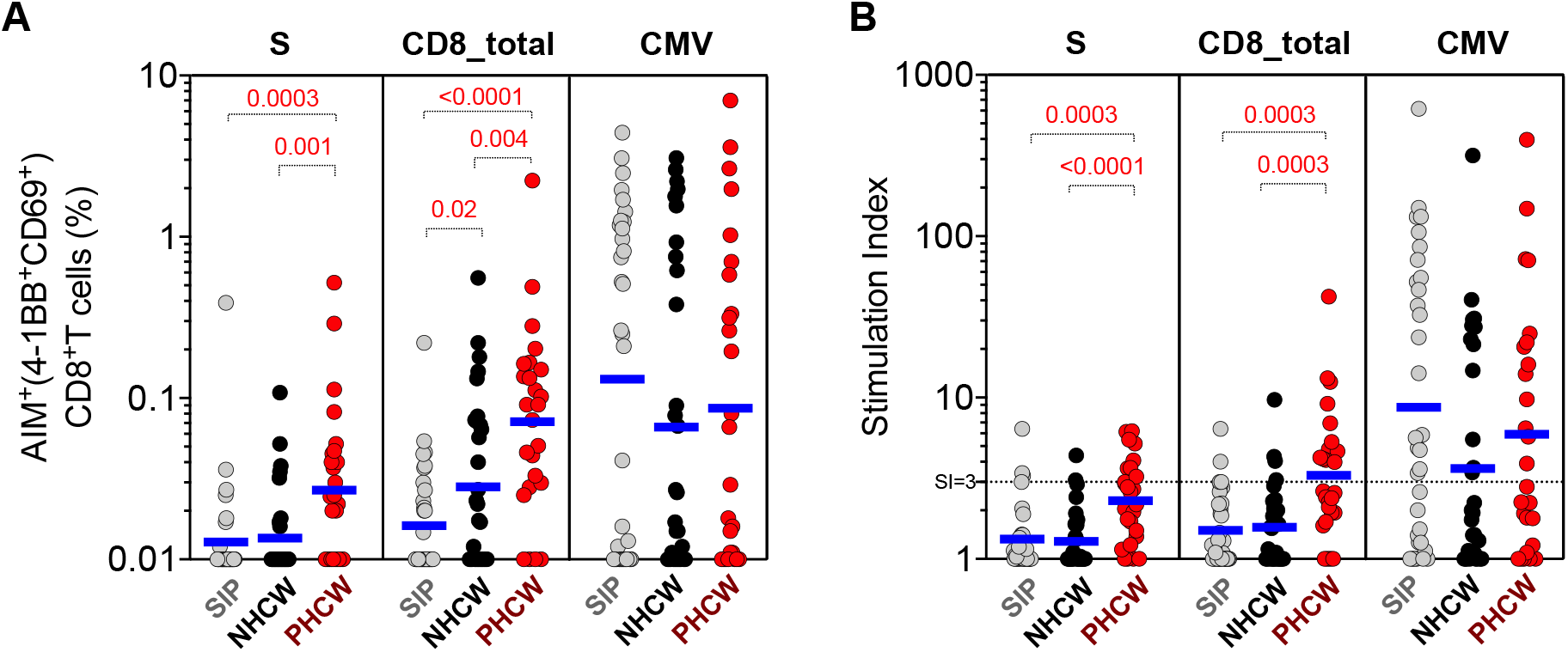
CD8+ T cell response to SARS-CoV-2 epitopes highest in PHCW and lowest in SIP. SARS-CoV-2-specific CD8+ T cells were measured as percentage of AIM+ (CD69+CD137+) CD8+ T cells after stimulation of PBMCs with spike only (“S”) MP or class I MPs (CD8A, CD8B). Graphs show data for specific responses against S, the combination of both CD8 MPs (CD8-total) and against CMV as a control, and plotted as (A) background subtracted or (B) as stimulation index (SI) against DMSO negative control. Geometric mean for the 3 different groups is shown. Non-parametric Kruskal-Wallis multiple comparison test was applied. P values are shown for the statistical significant comparisons. “SIP” = Shelter In Place community volunteers (n=33). “NHCW” = SeroNegative Health Care Workers (n=31). “PHCW” = Antibody or PCR Positive Health Care Workers (n=26).

In the case of the S pool, the CD8^+^ T cell response to SARS-CoV-2 spike protein was highest in PHCW (and similar between SIP and NHCW). More specifically, the total CD8^+^ T cell reactivity of the PHCW cohort to the SARS-CoV-2 pools was significantly higher than both NHCW (p=0.001 and p<0.0001 by the Kruskal-Wallis test for both absolute and SI readouts) and SIP (p=0.0003 and p=0.0003 for both absolute and SI readouts), as expected on the basis of the SARS-CoV-2 infection. The reactivity of the SIP and NHCW was not significantly different.

Similarly, with the CD8A+B pools, the total CD8^+^ T cell reactivity of the PHCW cohort to the SARS-CoV-2 pools was higher than both NHCW (p=0.004 and p=0.0003 for both absolute and SI readouts) and SIP (p <0.0001 and 0.0003 for both absolute and SI readouts). CD8^+^ T cell reactivity in all three cohorts was mediated by memory T cell subsets (**Supplementary Figure 5a-b)**. Furthermore, CD8^+^ T cell reactivity to SARS-CoV-2 peptides was associated with a trend towards an increased fraction of recently activated HLA-DR^+^CD38^+^ cells in the PHCW cohort (p=0.09), compared to the NHCW or SIP cohorts (**Supplementary Figure 5c)**. Overall these data suggest that the higher reactivity observed in NHCW as compared to SIP is largely confined to CD4^+^ T cell responses and only marginally seen in the case of CD8^+^ T cell responses, further suggesting that it is not resulting from infected individuals rapidly becoming seronegative.

## Discussion

Here we present evidence for differential reactivity to seasonal CCC and SARS-CoV-2 epitopes in different cohorts from the Miami metropolitan area. To the best of our knowledge, this is the first application of epitope pools to perform cross-sectional studies to investigate differential T cell reactivity to CCC and SARS-CoV-2 peptide sequences.

In particular, we present evidence that a cohort of HCW with presumed exposure to respiratory viruses is associated with higher levels of CCC reactive T cells as compared to a community SIP cohort, with presumed lower CCC exposure. Interestingly, similar CCC antibody levels were observed across all cohorts.

We hypothesized that this elevated level of CD4^+^ T cell reactivity was associated with higher reactivity against SARS-CoV-2 sequences, and indeed we show significantly higher levels of reactivity to SARS-CoV-2 sequences in the NHCW cohort. We also analyzed SARS-CoV-2 CD8^+^ T cell responses and observed a trend towards higher levels of SARS-CoV-2 cross-reactive CD8^+^ T cells in the NHCW compared to SIP controls. The detection of CD8^+^ T cell responses even in seronegative individuals is consistent with other studies (10, 12, 47), however, differences are less marked, consistent with lower levels of cross-reactivity reported for CD8^+^ T cells compared to CD4^+^ T cells (12, 14, 48).

While it is possible that some of the NHCW may have been infected in absence of seroconversion, or have been associated with transient seroconversion, we believe this is unlikely/infrequent. In fact, in our study we detected only 2 instances of transient seropositivity out of several hundreds who were studied. Based on this data and other published studies (23, 49) showing only a small fraction of convalescent individuals had borderline or absent IgG months after onset, we estimate that this scenario is uncommon, and the totality or vast majority of the seronegative donors has not been infected. Furthermore, our analysis found expression of cell markers associated with recent in vivo activation (11, 45) exclusively elevated in PHCW for both CD4^+^ and CD8^+^ T cell responses against SARS-CoV-2. As such, the patterns of reactivity detected in the NHCW are likely representative of a sampling of uninfected Miami HCW.

Samples from SARS-CoV-2 infected subjects were associated with lower levels of CCC reactivity as compared to non-exposed donors. This result was unexpected, but consistently detected in independent cohorts derived from Miami and San Diego. Several possibilities exist regarding the potential mechanisms underlying this effect. It is possible that SARS-CoV-2 infection may result in a generalized inhibition of CD4^+^ T cell responses to other CCCs but not unrelated viruses such as CMV. Impaired responses particularly associated with type I interferon activity in COVID-19 patients were also described in a recent report (50), suggesting that SARS-CoV-2 might interfere with innate immunity. SARS-CoV-2 infection may also result in expansion of SARS-CoV-2-specific, non-CCC reactive T cells, competing with the pre-existing CCC specificities (51, 52). Pre-existing CCC reactivity and different pre-exposure history can also influence disease severity and infection (28, 53). Based on our current understanding of viral dynamics, it appears unlikely that CD4^+^ T cells might be able to prevent infection, but it is possible that their presence may lead to rapid termination of infection and only transient seropositivity ((23, 54, 55) and see above) and reduce the ability of these individuals to infect others. It is also possible that CD8^+^ T cells might mediate or contribute to rapid termination of infection as described for SARS-CoV (56, 57) and other viral infection diseases (58-61). More recently, two studies reported that prior presumed CCC infections are associated with lower severity and reduced risk of COVID-19 disease (30, 62).

The present study builds on our previous study (15) that defined homology thresholds associated with CCC-SARS cross-reactivity, and allowed peptide pools expected to minimize such cross-reactivity. Indeed, some of the current challenges in identifying protective T cell responses against SARS-CoV-2 relates to distinguishing *bona fide* SARS-CoV-2 responses originating from SARS-CoV-2 infection from those associated with cross-reactive T cell memory generated by prior exposure to CCC. Further optimizations are in progress, based on inclusion of specific viral antigens and exclusion of specific epitopes. We anticipate that in the future we will be able to combine these improved pools, perhaps in conjunction with recently described whole blood assays (63, 64) to easily, accurately and independently assay levels of T cell reactivity to SARS-CoV-2 and CCC in different populations, such as pediatric cohorts versus older subjects, cohorts from different geographical locations associated with differential exposure to CCC, differential disease incidence, and other variables.

### Limitations of the present study

All recruited SARS-CoV-2 infected donors were associated with mild or asymptomatic disease, and therefore does not address another important aspect of research on CCC and SARS-CoV-2 interactions, namely whether levels of preexisting cross-reactive CCC T cell responses might influence disease severity (28, 30). Larger sample sizes will be required to analyze this issue in this type of cross-sectional design, but it is likely that a prospective longitudinal design might be necessary to firmly address this point based on evaluation of CCC reactivity in pre-infection samples, and its correlation with disease severity post SARS-CoV-2 infection. Additional limitations of this study are the relative small size of the cohorts investigated, and the unknown history of previous CCC exposure. Therefore, the results may not be necessarily generalizable to other situations with different patterns of prior exposure.

## Material and Methods

### Human subjects selection process

#### Sero<u>N</u>egative Health Care Workers (NHCW)

We observed relatively low prevalence of COVID-19 in HCW from the otolaryngology, ophthalmology, emergency medicine, and anesthesiology specialties at the University of Miami (https://coronavirus.miami.edu/dashboard/). These specialties either work closely to the face/airway and/or have frequent contact with patients with respiratory infections, in general, and, since the pandemic start, SARS-CoV-2. To investigate this matter further, we recruited individuals from these specialties **(Table 1)**.

The majority of high-risk HCW were physicians and nurses, though other support staff (e.g.: surgical/emergency room technicians) were enrolled as well, if they had sufficient occupational risk of contracting SARS-CoV-2. Individuals were included if they had a history of recent patient contact and no history of COVID-19 symptoms and no history of a positive PCR test or antibody testing. Blood draws from volunteer donors for serological studies were obtained in two separate sessions, the first on 4 April 2020 (Time Point 1) and the second about a month later, on 7-8 May 2020 (Time Point 2).

Despite significant opportunity for infection, the seropositivity rate for HCW was low. The seroprevalence rate at the first time-point was 1.05% (2/191). Both subjects affected had been asymptomatic since the start of the pandemic. Approximately one month later (7-8 May 2020) we re-tested individuals from the previous cohort as well as recruited new HCW to ascertain whether there had been a change in the seroconversion rate due to the growing intensity of the pandemic. At that time, 4.09% (9/220) were found to have seroconverted. Interestingly, the two participants who were seropositive during Time Point 1 no longer had detectable antibodies at Time Point 2.

Out of over 300 individuals who underwent antibody testing (Time point 1: n=191; Time point 2: n=220, 120 new and 100 repeat), larger blood donations were obtained from thirty-two individuals, for the purpose of isolating peripheral blood mononuclear cells (PBMCs) and analyzing T-cell responses. These individuals were selected on the basis of high patient contact, no history of a positive COVID-19 test or symptoms since the start of the pandemic, and recently confirmed seronegativity based on their participation in the Time Point 2 of the study. These blood donations were obtained from May 26 to June 3, 2020 (**Table 1**).

#### Antibody or PCR <u>P</u>ositive Health Care Workers (PHCW)

A cohort of HCWs working at the University of Miami Hospital with a history of patient contact and COVID-19 disease diagnosed by a positive antibody testing or positive nasopharyngeal swab PCR (*Positive Health Care Workers (PHCW))*, were included in this arm of the study (**Table 1**). These individuals gave blood donations for antibody testing and PBMC analysis. In addition, these subjects were asked about their COVID-19 symptoms and treatment course. PBMC samples collection after exposure ranged from 20 to 145 days (Median= 44 days) post-symptom onset (PSO). None had been admitted to the hospital for treatment for COVID-19 and were associated with mild (n=19) or asymptomatic disease (n=7). All these were SARS-CoV-2 positive by PCR (22/26) or antibody (4/26) testing. These collections took place from August 4 – 11, 2020. Effort was placed by the study team to balance this and the previous cohort in gender, age, and medical specialty **(Table 1)**.

#### Shelter In Place (SIP) Community Volunteers

To compare the high-risk HCW cohorts with community controls, individuals who had been sheltering in place during the pandemic with no connection to the health care industry (neither worked in the hospital nor had relatives/housemates that worked in the hospital) participated in the study. These individuals were acquaintances of HCW selected to represent the diverse ethnic communities of Miami. These individuals never had a positive PCR or antibody test nor had COVID-19 symptoms. Effort was placed by the study team to balance this and the previous cohorts in gender and age (**Table 1**). As above, these individuals gave blood donations for antibody testing and PBMC analysis. Sample collection took place in the June 4–16 2020 time-period.

#### Sero<u>N</u>egative (NSD) and Sero<u>P</u>ositive (COVID-19SD) donors from San Diego

In this study, two additional cohorts from San Diego were selected as validation cohorts. Individuals were selected on the basis of no history of a positive COVID-19 test and recently confirmed seronegativity (NSD) or with a history of COVID-19 disease diagnosed by a positive antibody testing (COVID-19SD) (**Table 1**).

Seronegative donors samples were obtained from local healthy unexposed adults in an anonymous fashion and protocols approved by the institutional review boards (IRB) of the La Jolla Institute (IRB#:VD-112).

Seropositive donors samples were either obtained at a UC San Diego Health clinic under the approved IRB protocols of the University of California, San Diego (UCSD; 200236X), recruited at the La Jolla Institute under IRB approved (LJI; VD-214) or provided by the CRO Sanguine that collected blood from previously PCR+ confirmed donors after resolution of symptoms. Subjects were asked about their COVID-19 symptoms. One donor has been admitted to the hospital for treatment for COVID-19 while the others were associated with mild (n=8) or moderate disease (n=1) (**Table 1**).

All collections took place from March-July, 2020 and blood was collected in acid citrate dextrose (ACD) tubes (UCSD) or in EDTA tubes (LJI and Sanguine) and stored at room temperature prior to processing for PBMC isolation and serum collection as described below.

### Peripheral blood mononuclear cells (PBMC) and serum isolation and handling

For the Miami cohorts, peripheral venous blood was collected in EDTA vacutainer tubes after obtaining written informed consent from the participants. PBMC were isolated by density gradient isolation using Ficoll-Paque (Lymphoprep, Nycomed Pharma, Oslo, Norway) as previously described and cryopreserved in 10% DMSO and 90% FCS in liquid nitrogen. Serum was collected and stored at −80°C. PBMC was shipped to LJI using dry shipper containing vapor phase liquid nitrogen and cryopreserved in liquid nitrogen until further use.

For the San Diego cohorts, whole blood was collected in heparin coated blood bags (healthy unexposed donors) or in ACD tubes (COVID-19 donors) and PBMCs isolated as described above and stored in liquid nitrogen until used in the assays. Serum samples were shipped in dry ice to Florian Lab at Icahn School of Medicine at Mount Sinai.

### OC43, NL63, HKU1, 229E and SARS-CoV-2 Enzyme-linked immunosorbent assay (ELISA)

Ninety-six-well microtiter plates (Thermo Fisher) were coated with 50 μL recombinant protein (OC43 spike, 229E spike, NL63 spike or HKU1 spike respectively) at a concentration of 2 ug/mL overnight at 4°C. The next day, the plates were washed three times with PBS (phosphate-buffered saline; Gibco) containing 0.1% Tween-20 (T-PBS, Fisher Scientific) using an automatic plate washer (BioTek). After washing, the plates were blocked for 1h at room temperature with 200 ul blocking solution (PBS-T supplemented with 3% (w/v) milk powder (American Bio)) per well. The blocking solution was thrown off and serum samples diluted to a starting concentration of 1:80, serially diluted 1:3 in PBS-T supplemented with 1% (w/v) milk powder and incubated at room temperature for 2 h. Using the automatic plate washer, the plates were washed three times with PBS-T and 50 ul anti-human IgG (Fab-specific) horseradish peroxidase antibody (HRP, Sigma, #A0293) diluted 1:3,000 in PBS-T (containing 1% milk powder) was added and incubated for 1 h at room temperature. The plates were washed three times and 100 μL SigmaFast o-phenylenediamine dihydrochloride (OPD; Sigma) was added to the wells. The enzymatic reaction was stopped after 10 minutes with 50 μL 3M hydrochloric acid (Thermo Fisher) per well and the plates read at a wavelength of 490 nm with a plate reader (Synergy H1, BioTek). The results were recorded in Microsoft Excel and the endpoint titers calculated using GraphPad Prism 8.

The SARS-CoV-2 ELISAs for both NHCW and PHCW were performed as previously described in detail (65) following a two-step ELISA protocol. In the first step serum samples were screened at a single serum dilution of 1:50 against the SARS-CoV-2 RBD. In the second step positive samples in the RBD ELISA were diluted to a starting concentration of 1:50 and serially diluted two-fold. The remainder of the protocol was performed similar as described above, with the exception that recombinant RBD or spike protein, respectively, were coated at a concentration of 2ug/mL. Alternatively, ELISA kits (Epitope Diagnostics, Inc (EDI™, San Diego, CA, USA) were used on the SIP community controls for COVID-19 IgM and IgG antibody detection to the N-antigen. The assays were performed according to manufacturer instructions with samples analyzed in duplicate with positive (PC), and negative (NC) controls using 100 µL (IgG) or 110 µL (IgM) of pre-diluted samples. Results were interpreted in accordance with the manufacturer’s cutoff calculations. A sample was determined to be positive if the OD reading was greater than the calculated positive cutoff value for that analytical batch. Testing was repeated if the quality control criteria was not met. Limits of detection were set at 1:80 and 1:50 for CCC and SARS-CoV-2 ELISA’s respectively. All data below was plotted as 1:25.

### Epitope predictions and peptide selection

We have previously predicted CD4^+^ and CD8^+^ T cell epitopes to study T cell responses with a high level of characterization for a number of studies, including allergies (66), tuberculosis (67), tetanus (34), pertussis (68) and DENV (61) utilizing the Immune Epitope Database and Analysis Resource (IEDB) (44). We have also previously developed the Megapool (MP) approach to allow simultaneous testing of large number of epitopes. According to this approach large numbers of different epitopes are solubilized, pooled and re-lyophilized to avoid cell toxicity problems associated with high concentrations of Dimethyl sulfoxide (DMSO) typically encountered when single pre-solubilized epitopes are pooled (43).

To investigate CCC CD4^+^ T cell responses, we performed prediction of peptides for HLA class II spanning the entire sequence of the 4 main CCC strains (HCoV-229E, HCoV-NL63, HCoV-HKU1 and HCoV-OC43). After selection of promiscuous binders, epitopes that shared 67% homology or more to SARS-CoV-2 sequences were removed from consideration and MPs composed of 15-mer peptides (overlapping by 10 amino acids) were generated and named low homology CCC pools (**Supplementary Table 1)**. The CMV MP is a pool of previously reported Class I and Class II epitopes (43). To study T cell responses against SARS-CoV-2, we used the entire SARS-CoV-2 genome (GenBank: MN908947) and we generated a MP of 15-mer peptides (overlapping by 10 amino acids) spanning the entire sequence of the spike protein and a MP for the remainder genome consistent of dominant HLA Class II predicted CD4^+^ T cell epitopes as previously described (44, 69). (**Supplementary Table 1)**.

Lastly, to measure CD8^+^ T cells responses against SARS-CoV-2, HLA Class I epitope prediction was performed as previously reported, using NetMHC pan EL 4.0 algorithm (70) for the top 12 more frequent HLA alleles and selecting the top 1 percentile predicted epitope per HLA allele clustered with nested/overlap reduction (12). The 628 predicted CD8^+^ T cell epitopes were split in two MPs containing 314 peptides each (CD8A and CD8B) (**Supplementary Table 1)**. We have previously shown that these MPs are suitable to stimulate T cell responses that are specific for SARS-CoV-2 from either exposed or non-exposed individuals, expanding the existing repertoire of T cell specificities (12, 14, 15, 39). All peptides were synthesized as crude material (A&A, San Diego, CA).

### Activation induced markers (AIM) assay and memory phenotype

Cryopreserved cells were thawed by diluting them in 10 ml complete RPMI 1640 with 5% human AB serum (Gemini Bioproducts) in the presence of benzonase [20ul/10mL], washed and stimulated for flow cytometry determinations. Specifically, activation induced cell marker (AIM) assays were performed as previously shown (34, 35) and cells cultured for 24 hours in the presence of CCC or SARS-CoV-2 specific MPs [1 μg/ml] in 96-wells U bottom plates at 1×10^6^ PBMC per well. A stimulation with an equimolar amount of DMSO was performed as negative control, phytohemagglutinin (PHA, Roche, 1μg/ml) and stimulation with a cytomegalovirus MP (CMV, 1μg/ml) were included as positive controls. Antibodies used in the AIM assay as well as the gating strategy used to define AIM reactive cells and memory sub-populations is listed in **Supplementary Table 2** and **Supplemental Figure 6**. AIM^+^ gates were drawn relative to the unstimulated condition for each donor. All samples were acquired on a ZE5 Cell analyzer (Bio-rad laboratories), and analyzed with FlowJo software (Tree Star, San Carlos, CA).

## Statistical analysis

Data and statistical analyses were done in FlowJo 10 and GraphPad Prism 8.4, unless otherwise stated. Data plotted in linear scale were expressed as Median. Data plotted in logarithmic scales were expressed as Geometric Mean. Non-parametric Mann-Whitney or Kruskal-Wallis test were applied for unpaired two-group or three-group comparisons, respectively. Correlation analysis were performed using non-parametric Spearman test. Details pertaining to significance are also noted in the respective legends and p<0.05 defined as statistical significant. All T cell data have been calculated as background subtracted data or stimulation index. Background subtracted data were derived by subtracting the percentage of AIM^+^ cells percentage after each MP stimulation from the DMSO stimulation. For all the DMSO values of 0 or lower than 0.005, DMSO was set to 0.005 and used for both subtracting and stimulation index analysis. When two stimuli were combined together, the percentage of AIM^+^ cells after SARS-CoV-2 stimulation was combined and either subtracted or divided by twice the value of the percentage of AIM+ cells derived from DMSO stimulation. All data below 0.01 or SI<1 were set to 0.01 or 1 for plotting and statistical analysis. Additional data analysis details are described in the respective figure legends.

## Author contributions

Conceptualization: S.G.P., M.E.H. and A.S. Formal analysis: R.dSA E.D.Y, S.P. and E.W. Funding acquisition: S.G.P. F.K. M.E.H and A.S. Investigation: R.dSA., S.P., E.W., E.D.Y., J.M., L.Q., D.S., K.J., F.A., and I.F. Resources: F.K., E.W., S.A.R., J.M.D, D.Ar., and D.An. Supervision: D.W., S.C, R.dSA., M.E.H., S.G.P, and A.S.; Writing: R.dSA, E.W., F.K., M.E.H., S.G.P. and A.S. R.dSA., S.P. and E.W. are equally first contributing authors for their shared efforts in the experimental work and investigation including clinical cohort coordination besides other described roles. S.G.P., M.E.H. and A.S are equally last contributing authors for their shared efforts in the supervision of the work besides the abovementioned roles. No gender or age bias were taken in consideration in the decision of the authors list order.

## Data Availability

No relevant data needs to be submitted to a public repository

## Acknowledgments

This work was funded by the NIH NIAID under awards AI142742 (Cooperative Centers for Human Immunology) (A.S., S.C.), NIH contract Nr. 75N9301900065 (D.W., A.S.), U01 AI141995-03 (A.S., B.P.), and U01 CA260541-01 (D.W). This work was partially supported by the NIAID Centers of Excellence for Influenza Research and Surveillance (CEIRS) contract HHSN272201400008C (FK, for reagent generation), Collaborative Influenza Vaccine Innovation Centers (CIVIC) contract 75N93019C00051 (FK, for reagent generation), NIH NIA R01 AG068110-01A1 (S.G.P, S.P), Miami CTSI emerging disease proposal (D.J., S.G.P, S.P.), University of Miami Institutional Support and the generous support of the JPB foundation, the Open Philanthropy Project (#2020-215611) and other philanthropic donations. We would like to thank Kizzmekia Corbett and Barney Graham at the Vaccine Research Center at NIH for providing plasmids for CCC spike expression. We wish to acknowledge all subjects for their participation and for donating their blood and time for this study. We thank the Clinical Core of the La Jolla Institute for Immunology and UCSD for sample collection and Allan Watson from Biorad for technical support with ZE5 Cell analyzer.

## Supplementary information for

**Supplementary Table 1a.** List of megapools (MP) used in this study

| MP name | Virus source | MP Type | Peptide number | Reference |
| --- | --- | --- | --- | --- |
| CMV | CMV | Predicted | 385 | Carrasco Pro et al., J Immun Res (2015) |
| 229E ≤ 60% | HCoV-229E | Predicted | 205 | Sup. Table 1b |
| HKU1 ≤ 60% | HCoV-HKU1 | Predicted | 272 | Sup. Table 1b |
| NL63 ≤ 60% | HCoV-NL63 | Predicted | 252 | Sup. Table 1b |
| OC43 ≤ 60% | HCoV-OC43 | Predicted | 258 | Sup. Table 1b |
| S | SARS-CoV-2 | Overlapping | 253 | Grifoni et al., Cell (2020) |
| CD4R | SARS-CoV-2 | Predicted | 221 | Grifoni et al., Cell (2020) |
| CD8A | SARS-CoV-2 | Predicted | 314 | Grifoni et al., Cell Host Microbe (2020) |
| CD8B | SARS-CoV-2 | Predicted | 314 | Grifoni et al., Cell Host Microbe (2020) |

**Supplementary Table 1b.** List of individual peptide sequences for each HCoV-CCC MP

| 229E (205 peptides) | HKU1 (272 peptides) | NL63 (252 peptides) | OC43 (258 peptides) |
| --- | --- | --- | --- |
| peptide 1 | peptide 1 | peptide 1 | peptide 1 |
| peptide 2 | peptide 2 | peptide 2 | peptide 2 |
| peptide 3 | peptide 3 | peptide 3 | peptide 3 |
| peptide ... | peptide ... | peptide ... | peptide ... |

**Supplementary Table 2.** List of antibodies used in the study

| Antibody | Fluorochrome | Clone | Vendor | Catalog number |
| --- | --- | --- | --- | --- |
| CD3 | AF700 | UCHT1 | eBiosciences | 56-0038-42 |
| CD4 | APCef780 | RPA-T4 | eBiosciences | 47-0049-42 |
| CD8 | BV650 | RPA-T8 | Biolegend | 301042 |
| CD137 | APC | 4B4-1 | Biolegend | 309810 |
| CD69 | PE | FN50 | BD Biosciences | 555531 |
| OX40 | PE-Cy7 | Ber-ACT35 | Biolegend | 350012 |
| CD38 | FITC | HB-7 | Biolegend | 356610 |
| HLA-DR | PE-CF594 | G46-6 | BD Biosciences | 562304 |
| CD45RA | eF450 | HI100 | Invitrogen | 48-0458-42 |
| CCR7 | PerCP/Cy5.5 | G043H7 | Biolegend | 353220 |
| CD14 | V500 | M5E2 | BD Biosciences | 561391 |
| CD19 | V500 | HIB19 | BD Biosciences | 561121 |
| Live/Dead Viability | eF506 | - | Invitrogen | 65-0866-18 |

**Supplementary Figure 1.**
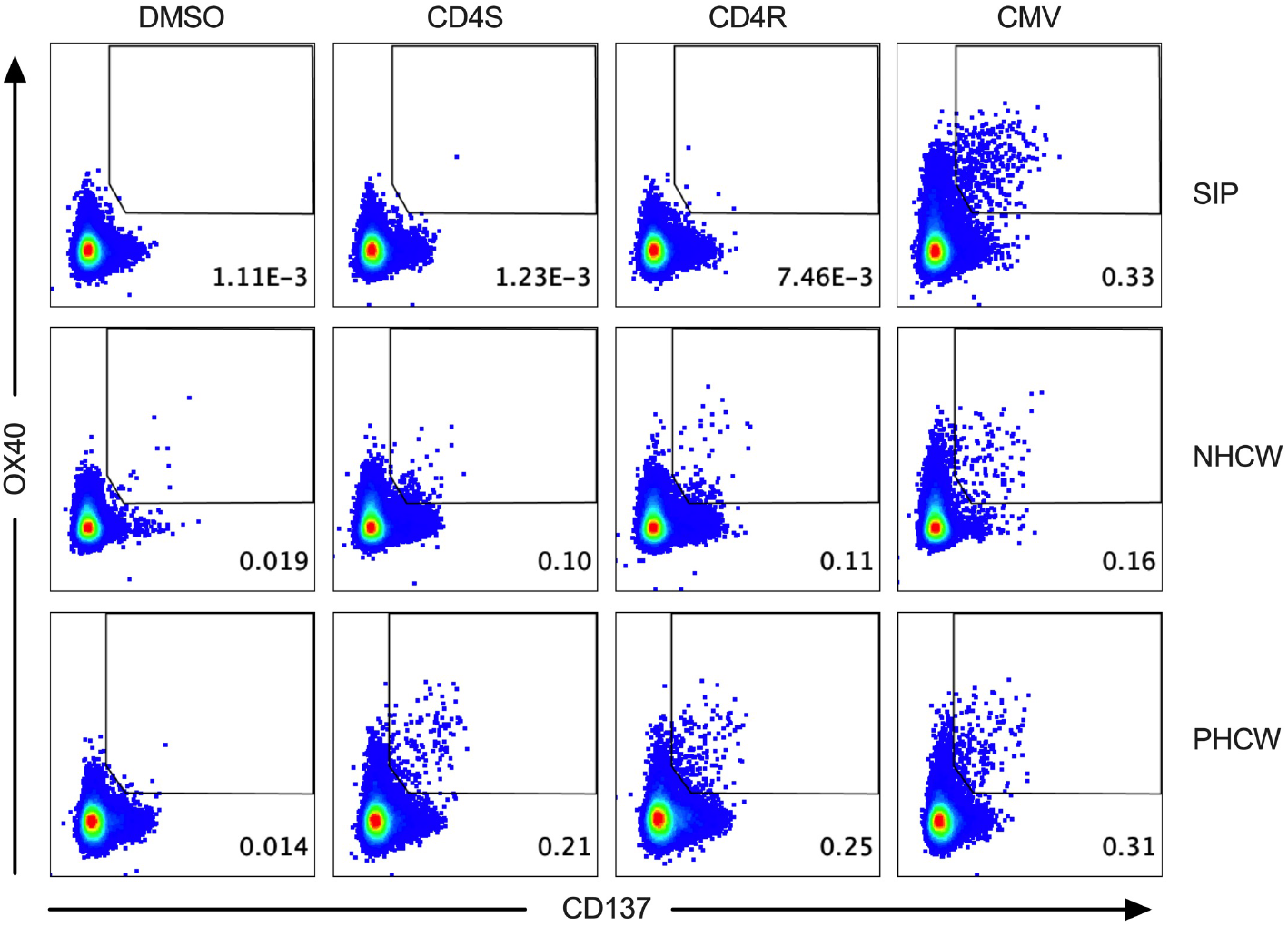
Representative flow cytometry gating of AIM+ (OX40/CD137) CD4+ T cell responses against SARS-CoV-2 and CMV across all the cohorts. Representative FACS plots, gated on total CD4+ T cells for the SARS-CoV-2-specific CD4+ T cells measured as percentage of AIM+ (OX40+CD137+) after stimulation of PBMCs with peptide pools encompassing spike (“S”) or the proteome without spike (“CD4R”) in addition to the DMSO negative control and the unrelated ubiquitous pathogen CMV across all the cohorts. Cell frequency for AIM+ cells in the several conditions is indicated

**Supplementary Figure 2.**
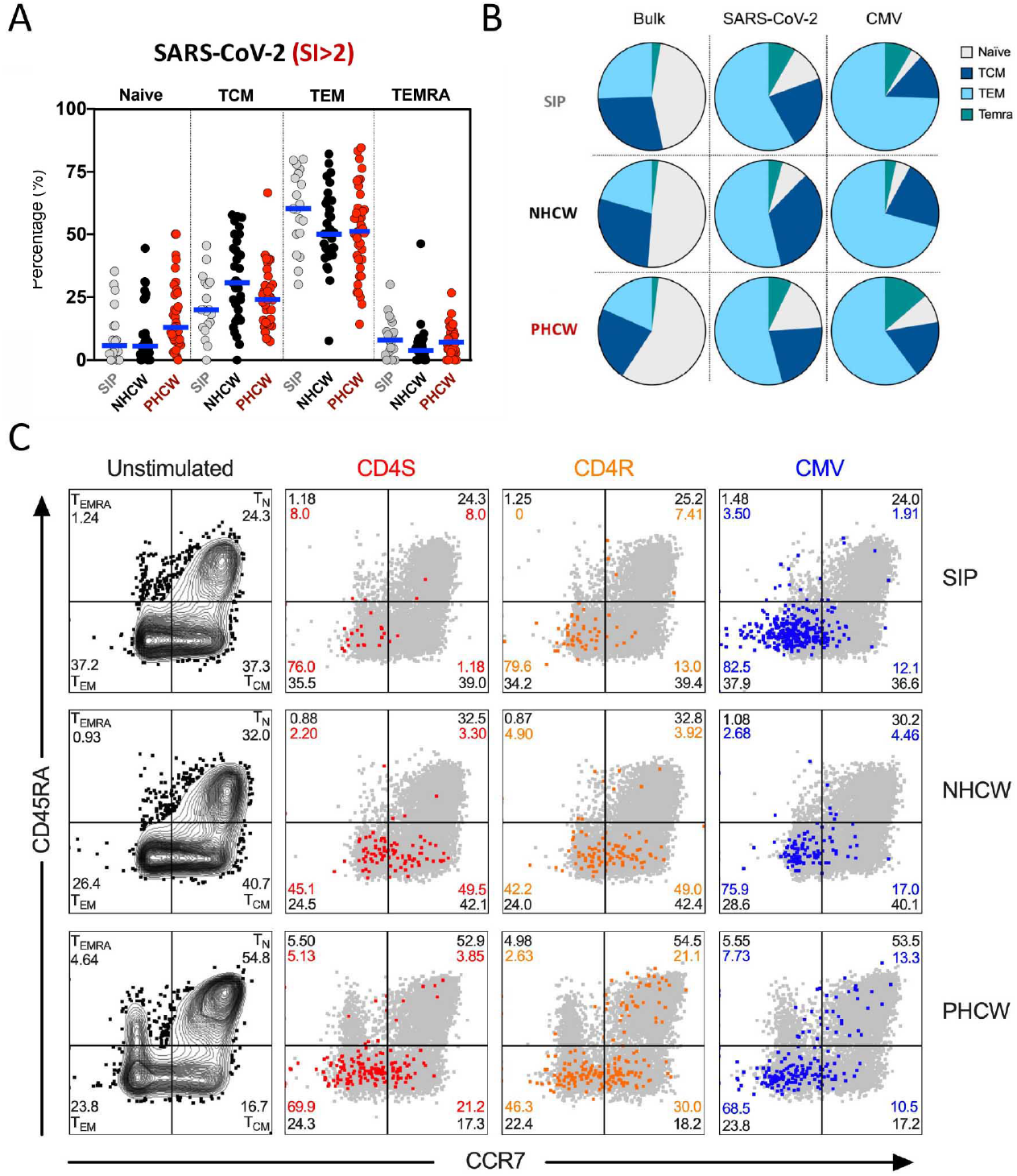
CD4+ T cell reactivity to SARS-CoV-2 epitopes is mediated by memory cells. SARS-CoV-2-specific CD4+ T cell subsets (Tn: CD45RA+ CCR7+, Temra: CD45RA+ CCR7-, Tcm: CD45RA-CCR7+ and Tem: CD45RA-CCR7-) were measured after stimulation of PBMCs with peptide pools encompassing spike (“S”) or the proteome without spike (“CD4R”). (A) Phenotype of antigen-specific CD4+ T cells (OX40+CD137+) responding to the indicated pools of SARS-CoV-2 and with SI>2 in each cohort. Each dot represents the response of an individual subject to an individual pool and geometric mean for the 3 different groups is shown (B) Overall averages for total CD4+ (Bulk) or antigen-specific CD4+ T cell subsets for SARS-CoV-2 or CMV detected in the 3 different cohorts. (C) Representative FACS plots, gated on total CD4+ T cells (grey) for the SARS-CoV-2-specific CD4+ T cells (colored) measured as percentage of AIM+ (OX40+CD137+) after stimulation of PBMCs with peptide pools encompassing spike (“S”) or the proteome without spike (“CD4R”) in addition to the DMSO negative control and the unrelated ubiquitous pathogen CMV across all the cohorts. Cell frequency for each of the subsets is indicated in each quadrant for AIM+ cells (colored) or total (“bulk”) CD4+ T cells (black).

**Supplementary Figure 3.**
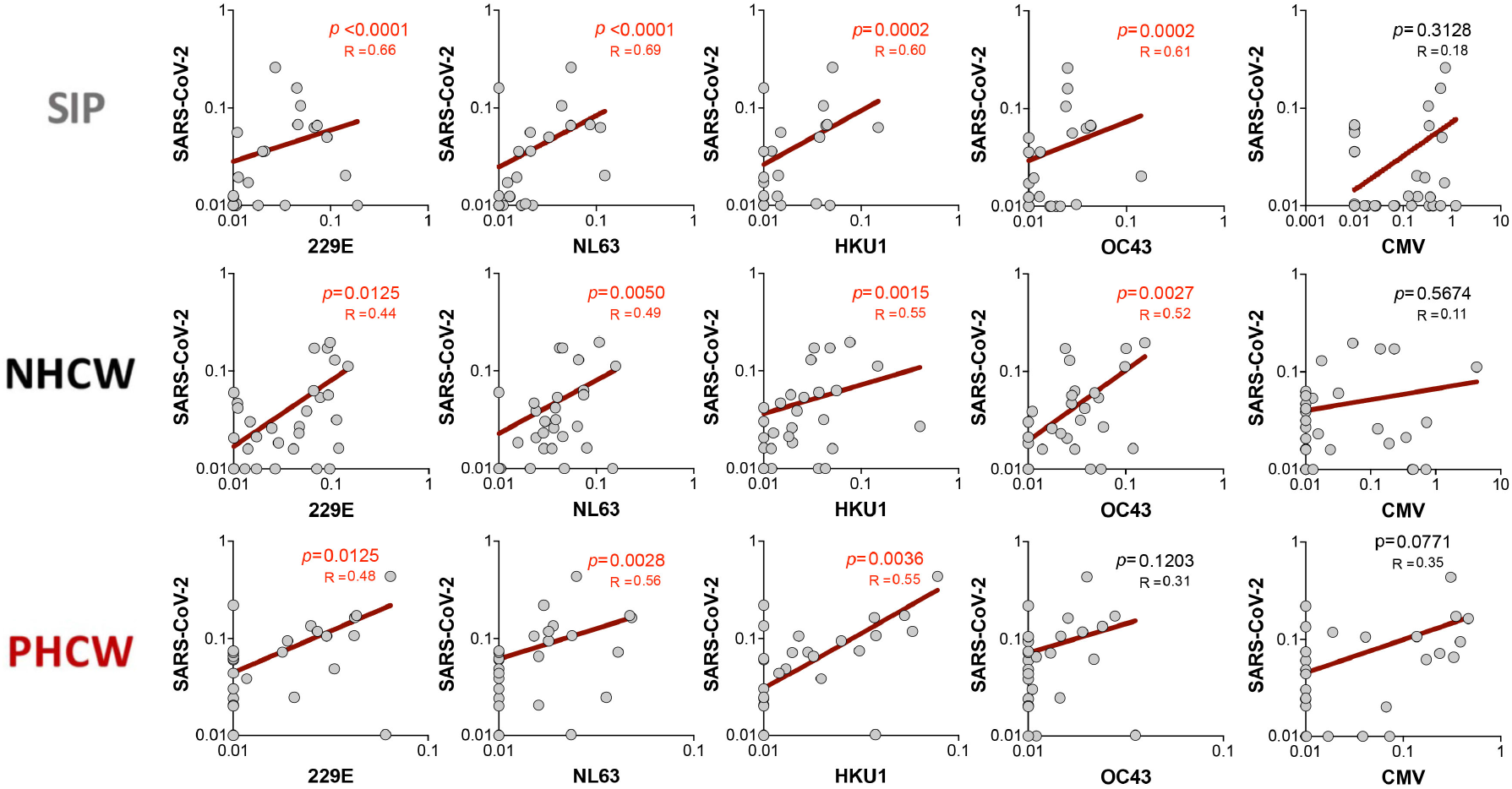
Correlation of SIP, NHCW and PHCW cohort responses to CCC and SARS-CoV-2. Correlation between SARS-CoV-2-specific CD4+ T cells and each of the 4 CCC-specific CD4+ T cells (HCoV-229E, HCoV-NL63, HCoV-HKU1 and HCoV-OC43) as well as CMV. Total SARS-CoV-2 MP responses per donor were used in each case (“Spike” + “Non-spike” (CD4-total). Statistical comparisons were performed using Spearman correlations. Non-linear fit curve and P and R values are shown.

**Supplementary Figure 4.**
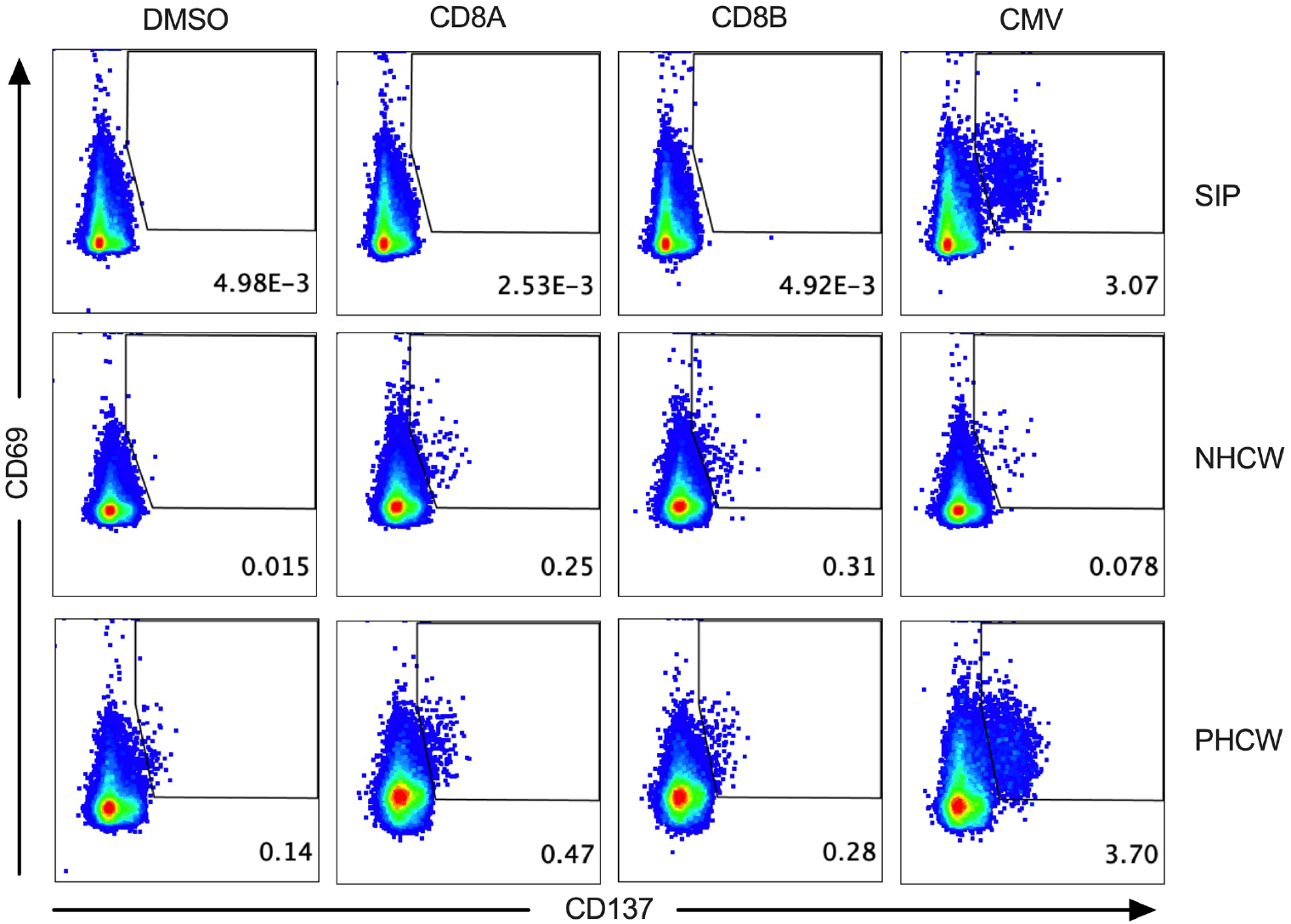
Representative flow cytometry gating of AIM+ (CD69/CD137) CD8+ T cell responses against SARS-CoV-2 and CMV across all the cohorts. Representative FACS plots, gated on total CD8+ T cells for the SARS-CoV-2-specific CD8+ T cells measured as percentage of AIM+ (CD69+CD137+) after stimulation of PBMCs with class I MPs (CD8A, CD8B) in addition to the DMSO negative control and the unrelated ubiquitous pathogen CMV across all the cohorts. Cell frequency for AIM+ cells in the several conditions is indicated

**Supplementary Figure 5.**
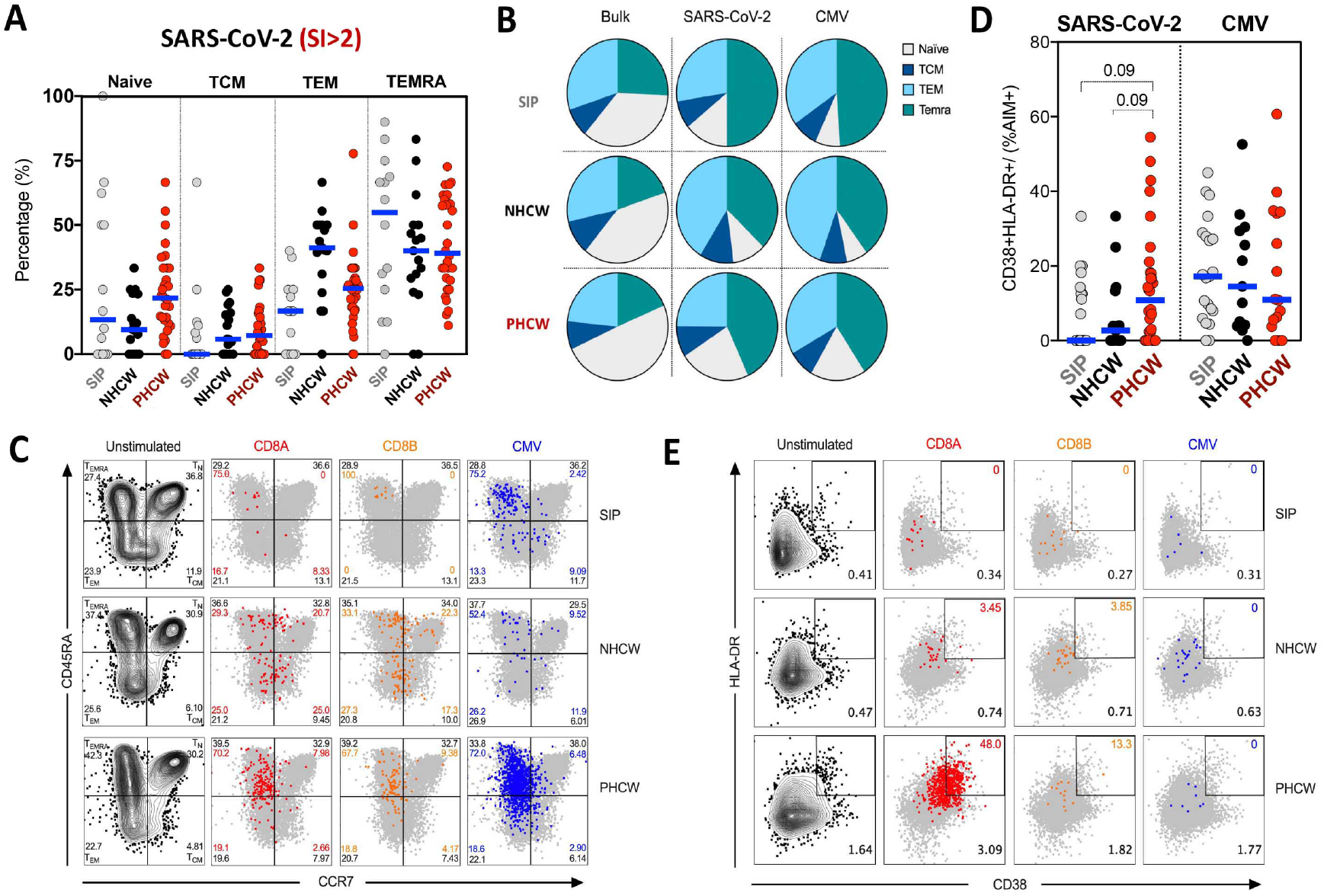
CD8+ T cell reactivity to SARS-CoV-2 epitopes is mediated by memory cells and associated with recent infection in PHCW. SARS-CoV-2-specific CD8+ T cell subsets (Tn: CD45RA+ CCR7+, Temra: CD45RA+ CCR7-, Tcm: CD45RA-CCR7+ and Tem: CD45RA-CCR7-) were measured after stimulation of PBMCs with class I MPs (CD8A, CD8B). (A) Phenotype of antigen-specific CD4+ T cells (CD69+CD137+) responding to the indicated pools of SARS-CoV-2 and with SI>2 in each cohort. Each dot represents the response of an individual subject to an individual pool and geometric mean for the 3 different groups is shown (B) Overall averages for total CD8+ (Bulk) or antigen-specific CD8+ T cell subsets for SARS-CoV-2 or CMV detected in the 3 different cohorts. (C) Representative FACS plots, gated on total CD8+ T cells (grey) for the SARS-CoV-2-specific CD8+ T cells (colored) measured as percentage of AIM+ (CD69+CD137+) after stimulation of PBMCs with class I MPs (CD8A, CD8B) in addition to the DMSO negative control and the unrelated ubiquitous pathogen CMV across all the cohorts. Cell frequency for each of the subsets is indicated in each quadrant for AIM+ cells (colored) or total (“bulk”) CD4+ T cells (black). (D) Recently activated SARS-CoV-2-specific CD8+ T cells were measured as percentage of CD38+/HLA-DR+ cells in AIM+ (CD69+CD137+) CD8+ T cells after stimulation of PBMCs with class I MPs (CD8A, CD8B). Graphs show data for specific responses against SARS-CoV-2 and CMV responses with SI>2. Each dot represents the response of an individual subject to an individual pool. Geometric mean for the 3 different groups is shown. Non-parametric Kruskal-Wallis multiple comparison test was applied. P values are shown with p<0.05 defined as statistical significant. (E) Representative FACS plots of HLA-DR/CD38+ cells in AIM+ (CD69+CD137+) CD8+ T cells (colored) overlapped with total HLA-DR/CD38 expression (grey) for all the cohorts in the different unstimulated or stimulated conditions. Cell frequency of HLA-DR/CD38+ in AIM+ cells or total CD8+ T cells is indicated on the top and bottom right corner respectively.

**Supplementary Figure 6.**
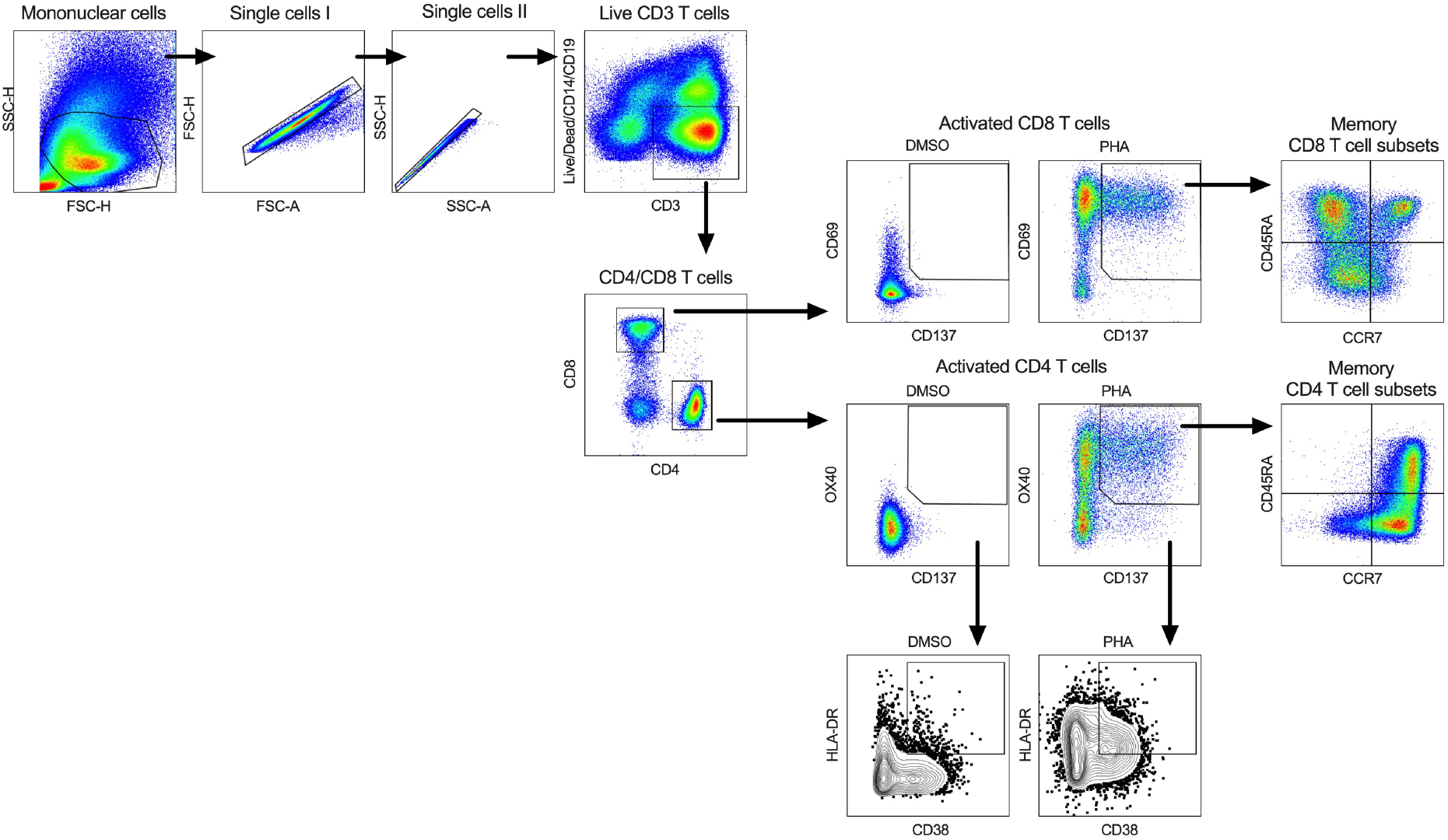
Gating strategy for CD4+ T cell and CD8+ T cell AIM assays used in this study. Example of flow cytometry gating strategy. Briefly, mononuclear cells were gated out of all events followed by subsequent singlet gating. Live CD3+ T cells were gated out from dead cells, B cells and Monocytes. T cells were then divided as CD4+ or CD8+ and each population further subdivided into either AIM_CD4+ (OX40/CD137) or AIM_CD8+ (CD69/CD137) respectively or alternatively as HLA-DR/CD38. AIM+ cells were further mapped into memory subsets as a function of CCR7 and CD45RA expression (Tn: CD45RA+ CCR7+, Temra: CD45RA+ CCR7-, Tcm: CD45RA-CCR7+ and Tem: CD45RA-CCR7-).

## References

1. Nguyen LH, Drew DA, Graham MS, Joshi AD, Guo CG, Ma W, et al. Risk of COVID-19 among front-line health-care workers and the general community: a prospective cohort study. Lancet Public Health. 2020;5(9):e475–e83.

2. Iversen K, Bundgaard H, Hasselbalch RB, Kristensen JH, Nielsen PB, Pries-Heje M, et al. Risk of COVID-19 in health-care workers in Denmark: an observational cohort study. Lancet Infect Dis. 2020.

3. Moscola J, Sembajwe G, Jarrett M, Farber B, Chang T, McGinn T, et al. Prevalence of SARS-CoV-2 Antibodies in Health Care Personnel in the New York City Area. JAMA. 2020;324(9):893–5.

4. Rudberg AS, Havervall S, Manberg A, Jernbom Falk A, Aguilera K, Ng H, et al. SARS-CoV-2 exposure, symptoms and seroprevalence in healthcare workers in Sweden. Nat Commun. 2020;11(1):5064.

5. Zhang S, Guo M, Wu F, Xiong N, Ma Y, Wang Z, et al. Factors associated with asymptomatic infection in health-care workers with severe acute respiratory syndrome coronavirus 2 infection in Wuhan, China: a multicentre retrospective cohort study. Clin Microbiol Infect. 2020.

6. Venugopal U, Jilani N, Rabah S, Shariff MA, Jawed M, Batres AM, et al. SARS-CoV-2 Seroprevalence Among Health Care Workers in a New York City Hospital: A Cross-Sectional Analysis During the COVID-19 Pandemic. Int J Infect Dis. 2020.

7. Reynolds CJ, Swadling L, Gibbons JM, Pade C, Jensen MP, …., et al. Healthcare workers with mild / asymptomatic SARS-CoV-2 infection show T cell responses and neutralising antibodies after the first wave. Preprint at doiorg/101101/2020101320211763. 2020.

8. Vallejo A, Vizcarra P, Quereda C, Moreno A, and Casado JL. SARS-CoV-2 cellular immune responses in uninfected health care workers. Preprint at https://1021203/rs3rs-55720/v1. 2020.

9. Sotgiu G, Barassi A, Miozzo M, Saderi L, Piana A, Orfeo N, et al. SARS-CoV-2 specific serological pattern in healthcare workers of an Italian COVID-19 forefront hospital. BMC Pulm Med. 2020;20(1):203.

10. Le Bert N, Tan AT, Kunasegaran K, Tham CYL, Hafezi M, Chia A, et al. SARS-CoV-2-specific T cell immunity in cases of COVID-19 and SARS, and uninfected controls. Nature. 2020;584(7821):457–62.

11. Sekine T, Perez-Potti A, Rivera-Ballesteros O, Stralin K, Gorin JB, Olsson A, et al. Robust T Cell Immunity in Convalescent Individuals with Asymptomatic or Mild COVID-19. Cell. 2020;183(1):158–68 e14.

12. Grifoni A, Weiskopf D, Ramirez SI, Mateus J, Dan JM, Moderbacher CR, et al. Targets of T Cell Responses to SARS-CoV-2 Coronavirus in Humans with COVID-19 Disease and Unexposed Individuals. Cell. 2020;181(7):1489–501 e15.

13. Braun J, Loyal L, Frentsch M, Wendisch D, Georg P, Kurth F, et al. SARS-CoV-2-reactive T cells in healthy donors and patients with COVID-19. Nature. 2020.

14. Rydyznski Moderbacher C, Ramirez SI, Dan JM, Grifoni A, Hastie KM, Weiskopf D, et al. Antigen-Specific Adaptive Immunity to SARS-CoV-2 in Acute COVID-19 and Associations with Age and Disease Severity. Cell. 2020.

15. Mateus J, Grifoni A, Tarke A, Sidney J, Ramirez SI, Dan JM, et al. Selective and cross-reactive SARS-CoV-2 T cell epitopes in unexposed humans. Science. 2020.

16. English KM, Langley JM, McGeer A, Hupert N, Tellier R, Henry B, et al. Contact among healthcare workers in the hospital setting: developing the evidence base for innovative approaches to infection control. BMC Infect Dis. 2018;18(1):184.

17. Chughtai AA, Stelzer-Braid S, Rawlinson W, Pontivivo G, Wang Q, Pan Y, et al. Contamination by respiratory viruses on outer surface of medical masks used by hospital healthcare workers. BMC Infect Dis. 2019;19(1):491.

18. Jacobs JL, Ohde S, Takahashi O, Tokuda Y, Omata F, and Fukui T. Use of surgical face masks to reduce the incidence of the common cold among health care workers in Japan: a randomized controlled trial. Am J Infect Control. 2009;37(5):417–9.

19. Killerby ME, Biggs HM, Haynes A, Dahl RM, Mustaquim D, Gerber SI, et al. Human coronavirus circulation in the United States 2014-2017. J Clin Virol. 2018;101:52–6.

20. Nickbakhsh S, Ho A, Marques DFP, McMenamin J, Gunson RN, and Murcia PR. Epidemiology of Seasonal Coronaviruses: Establishing the Context for the Emergence of Coronavirus Disease 2019. J Infect Dis. 2020;222(1):17–25.

21. Cimolai N. Complicating Infections Associated with Common Endemic Human Respiratory Coronaviruses. Health Secur. 2020.

22. Mateus J, Grifoni A, Tarke A, Sidney J, Ramirez SI, Dan JM, et al. Selective and cross-reactive SARS-CoV-2 T cell epitopes in unexposed humans. Science. 2020;370(6512):89–94.

23. Nelde A, Bilich T, Heitmann JS, Maringer Y, Salih HR, Roerden M, et al. SARS-CoV-2-derived peptides define heterologous and COVID-19-induced T cell recognition. Nat Immunol. 2020.

24. Klompus S, Leviatan S, Vogl T, Kalka I, Godneva A, Shinar E, et al. Cross-reactive antibody responses against SARS-CoV-2 and seasonal common cold coronaviruses. Preprint at https://doiorg/101101/2020090120182220. 2020.

25. Henss L, Scholz T, von Rhein C, Wieters I, Borgans F, Eberhardt FJ, et al. Analysis of humoral immune responses in SARS-CoV-2 infected patients. J Infect Dis. 2020.

26. Aydillo T, Rombauts A, Stadlbauer D, Aslam S, Abelenda-Alonso G, Escalera A, et al. Antibody Immunological Imprinting on COVID-19 Patients. Preprint at https://doiorg/101101/2020101420212662. 2020.

27. Ng KW, Faulkner N, Cornish GH, Rosa A, Harvey R, Hussain S, et al. Preexisting and de novo humoral immunity to SARS-CoV-2 in humans. Science. 2020;370(6522):1339–43.

28. Lipsitch M, Grad YH, Sette A, and Crotty S. Cross-reactive memory T cells and herd immunity to SARS-CoV-2. Nat Rev Immunol. 2020.

29. Meyerholz DK, and Perlman S. Does common cold coronavirus infection protect against severe SARS-CoV2 disease? J Clin Invest. 2020.

30. Sagar M, Reifler K, Rossi M, Miller NS, Sinha P, White L, et al. Recent endemic coronavirus infection is associated with less severe COVID-19. J Clin Invest. 2020.

31. Wyllie D, Mulchandani R, Hayley E Jones, Taylor-Phillips S, BrooksAndre T, Ae C, et al. SARS-CoV-2 responsive T cell numbers are associated with protection from COVID-19: A prospective cohort study in keyworkers. Preprint at https://doiorg101101/2020110220222778;. 2020.

32. Beretta A, Cranage M, and Zipeto D. Is Cross-Reactive Immunity Triggering COVID-19 Immunopathogenesis? Front Immunol. 2020;11:567710.

33. Edridge AWD, Kaczorowska J, Hoste ACR, Bakker M, Klein M, Loens K, et al. Seasonal coronavirus protective immunity is short-lasting. Nat Med. 2020.

34. da Silva Antunes R, Paul S, Sidney J, Weiskopf D, Dan JM, Phillips E, et al. Definition of Human Epitopes Recognized in Tetanus Toxoid and Development of an Assay Strategy to Detect Ex Vivo Tetanus CD4+ T Cell Responses. PloS one. 2017;12(1):e0169086.

35. Dan JM, Lindestam Arlehamn CS, Weiskopf D, da Silva Antunes R, Havenar-Daughton C, Reiss SM, et al. A Cytokine-Independent Approach To Identify Antigen-Specific Human Germinal Center T Follicular Helper Cells and Rare Antigen-Specific CD4+ T Cells in Blood. Journal of immunology. 2016;197(3):983–93.

36. da Silva Antunes R, Babor M, Carpenter C, Khalil N, Cortese M, Mentzer AJ, et al. Th1/Th17 polarization persists following whole-cell pertussis vaccination despite repeated acellular boosters. J Clin Invest. 2018;128(9):3853–65.

37. Reiss S, Baxter AE, Cirelli KM, Dan JM, Morou A, Daigneault A, et al. Comparative analysis of activation induced marker (AIM) assays for sensitive identification of antigen-specific CD4 T cells. PloS one. 2017;12(10):e0186998.

38. Voic H, de Vries RD, Sidney J, Rubiro P, Moore E, Phillips E, et al. Identification and characterization of CD4+ T cell epitopes after Shingrix vaccination. J Virol. 2020.

39. Weiskopf D, Schmitz KS, Raadsen MP, Grifoni A, Okba NMA, Endeman H, et al. Phenotype and kinetics of SARS-CoV-2-specific T cells in COVID-19 patients with acute respiratory distress syndrome. Sci Immunol. 2020;5(48).

40. Paul S, Lindestam Arlehamn CS, Scriba TJ, Dillon MB, Oseroff C, Hinz D, et al. Development and validation of a broad scheme for prediction of HLA class II restricted T cell epitopes. J Immunol Methods. 2015;422:28–34.

41. Dhanda SK, Vaughan K, Schulten V, Grifoni A, Weiskopf D, Sidney J, et al. Development of a novel clustering tool for linear peptide sequences. Immunology. 2018;155(3):331–45.

42. Weiskopf D, Bangs DJ, Sidney J, Kolla RV, De Silva AD, de Silva AM, et al. Dengue virus infection elicits highly polarized CX3CR1+ cytotoxic CD4+ T cells associated with protective immunity. Proceedings of the National Academy of Sciences of the United States of America. 2015;112(31):E4256–63.

43. Carrasco Pro S, Sidney J, Paul S, Lindestam Arlehamn C, Weiskopf D, Peters B, et al. Automatic Generation of Validated Specific Epitope Sets. Journal of immunology research. 2015;2015:763461.

44. Grifoni A, Sidney J, Zhang Y, Scheuermann RH, Peters B, and Sette A. A Sequence Homology and Bioinformatic Approach Can Predict Candidate Targets for Immune Responses to SARS-CoV-2. Cell Host Microbe. 2020;27(4):671–80 e2.

45. Kuri-Cervantes L, Pampena MB, Meng W, Rosenfeld AM, Ittner CAG, Weisman AR, et al. Comprehensive mapping of immune perturbations associated with severe COVID-19. Sci Immunol. 2020;5(49).

46. Mathew D, Giles JR, Baxter AE, Oldridge DA, Greenplate AR, Wu JE, et al. Deep immune profiling of COVID-19 patients reveals distinct immunotypes with therapeutic implications. Science. 2020;369(6508).

47. Schulien I, Kemming J, Oberhardt V, Wild K, Seidel LM, Killmer S, et al. Characterization of preexisting and induced SARS-CoV-2-specific CD8(+) T cells. Nat Med. 2020.

48. Ferretti AP, Kula T, Wang Y, Nguyen DMV, Weinheimer A, Dunlap GS, et al. Unbiased Screens Show CD8(+) T Cells of COVID-19 Patients Recognize Shared Epitopes in SARS-CoV-2 that Largely Reside outside the Spike Protein. Immunity. 2020;53(5):1095–107 e3.

49. Schwarzkopf S, Krawczyk A, Knop D, Klump H, Heinold A, Heinemann FM, et al. Cellular Immunity in COVID-19 Convalescents with PCR-Confirmed Infection but with Undetectable SARS-CoV-2-Specific IgG. Emerg Infect Dis. 2020;27(1).

50. Hadjadj J, Yatim N, Barnabei L, Corneau A, Boussier J, Smith N, et al. Impaired type I interferon activity and inflammatory responses in severe COVID-19 patients. Science. 2020;369(6504):718–24.

51. Bacher P, Rosati E, Esser D, Martini GR, Saggau C, Schiminsky E, et al. Low-Avidity CD4(+) T Cell Responses to SARS-CoV-2 in Unexposed Individuals and Humans with Severe COVID-19. Immunity. 2020;53(6):1258–71 e5.

52. Tarke A, Sidney J, Kidd CK, Dan JM, Ramirez SI, Yu ED, et al. Comprehensive analysis of T cell immunodominance and immunoprevalence of SARS-CoV-2 epitopes in COVID-19 cases. Preprint at https://doiorg/101101/20201208416750. 2020.

53. de Vries RD. SARS-CoV-2-specific T-cells in unexposed humans: presence of cross-reactive memory cells does not equal protective immunity. Signal Transduct Target Ther. 2020;5(1):224.

54. Elong Ngono A, Young MP, Bunz M, Xu Z, Hattakam S, Vizcarra E, et al. CD4+ T cells promote humoral immunity and viral control during Zika virus infection. PLoS Pathog. 2019;15(1):e1007474.

55. Walton S, Mandaric S, and Oxenius A. CD4 T cell responses in latent and chronic viral infections. Front Immunol. 2013;4:105.

56. Channappanavar R, Fett C, Zhao J, Meyerholz DK, and Perlman S. Virus-specific memory CD8 T cells provide substantial protection from lethal severe acute respiratory syndrome coronavirus infection. J Virol. 2014;88(19):11034–44.

57. Chen H, Hou J, Jiang X, Ma S, Meng M, Wang B, et al. Response of memory CD8+ T cells to severe acute respiratory syndrome (SARS) coronavirus in recovered SARS patients and healthy individuals. Journal of immunology. 2005;175(1):591–8.

58. Hemann EA, Kang SM, and Legge KL. Protective CD8 T cell-mediated immunity against influenza A virus infection following influenza virus-like particle vaccination. Journal of immunology. 2013;191(5):2486–94.

59. Schmidt ME, and Varga SM. The CD8 T Cell Response to Respiratory Virus Infections. Front Immunol. 2018;9:678.

60. Thimme R, Wieland S, Steiger C, Ghrayeb J, Reimann KA, Purcell RH, et al. CD8(+) T cells mediate viral clearance and disease pathogenesis during acute hepatitis B virus infection. J Virol. 2003;77(1):68–76.

61. Elong Ngono A, Chen HW, Tang WW, Joo Y, King K, Weiskopf D, et al. Protective Role of Cross-Reactive CD8 T Cells Against Dengue Virus Infection. EBioMedicine. 2016;13:284–93.

62. Aran D, Beachler DC, Lanes S, and Overhage JM. Prior Presumed Coronavirus Infection Reduces COVID-19 Risk: A Cohort Study. J Infect. 2020.

63. Murugesan K, Jagannathan P, Pham TD, Pandey S, Bonilla HF, Jacobson K, et al. Interferon-gamma release assay for accurate detection of SARS-CoV-2 T cell response. Clin Infect Dis. 2020.

64. Petrone L, Petruccioli E, Vanini V, Cuzzi G, Najafi Fard S, Alonzi T, et al. A whole blood test to measure SARS-CoV-2-specific response in COVID-19 patients. Clin Microbiol Infect. 2020.

65. Stadlbauer D, Amanat F, Chromikova V, Jiang K, Strohmeier S, Arunkumar GA, et al. SARS-CoV-2 Seroconversion in Humans: A Detailed Protocol for a Serological Assay, Antigen Production, and Test Setup. Curr Protoc Microbiol. 2020;57(1):e100.

66. Hinz D, Seumois G, Gholami AM, Greenbaum JA, Lane J, White B, et al. Lack of allergy to timothy grass pollen is not a passive phenomenon but associated with the allergen-specific modulation of immune reactivity. Clinical and experimental allergy: journal of the British Society for Allergy and Clinical Immunology. 2016;46(5):705–19.

67. Lindestam Arlehamn CS, McKinney DM, Carpenter C, Paul S, Rozot V, Makgotlho E, et al. A Quantitative Analysis of Complexity of Human Pathogen-Specific CD4 T Cell Responses in Healthy M. tuberculosis Infected South Africans. PLoS Pathog. 2016;12(7):e1005760.

68. Bancroft T, Dillon MB, da Silva Antunes R, Paul S, Peters B, Crotty S, et al. Th1 versus Th2 T cell polarization by whole-cell and acellular childhood pertussis vaccines persists upon re-immunization in adolescence and adulthood. Cellular immunology. 2016;304-305:35–43.

69. Grifoni A, Voic H, Dhanda SK, Kidd CK, Brien JD, Buus S, et al. T Cell Responses Induced by Attenuated Flavivirus Vaccination Are Specific and Show Limited Cross-Reactivity with Other Flavivirus Species. J Virol. 2020;94(10).

70. Jurtz V, Paul S, Andreatta M, Marcatili P, Peters B, and Nielsen M. NetMHCpan-4.0: Improved Peptide-MHC Class I Interaction Predictions Integrating Eluted Ligand and Peptide Binding Affinity Data. Journal of immunology. 2017;199(9):3360–8.

